# Revisiting the UK Genetic Severity Score for NF2: a proposal for adding functional information

**DOI:** 10.1101/2020.10.22.20216614

**Authors:** N. Catasús, B. García, I. Galvan, A. Plana, A. Negro, I. Rosas, A. Ros, E. Amilibia, JL. Becerra, C. Hostalot, F. Roca-Ribas, I. Bielsa, C. Lazaro, R. de Cid, E. Serra, I. Blanco, E. Castellanos, on behalf of the NF2 Multidisciplinary Clinics HUGTiP-ICO-IMPPC

## Abstract

**Background:** Neurofibromatosis Type 2 (NF2) is an autosomal dominant disorder characterized by the development of multiple schwannomas, particularly at vestibular nerves, and meningiomas. The UK NF2 Genetic Severity Score (GSS) helps predicting the disease course from germline *NF2* pathogenic variants, optimizing the clinical follow-up and the genetic counselling offered to affected families.

**Methods:** Spanish Reference Center patients were classified following the GSS and patients’ clinical severity was measured and compared between GSS groups. The GSS was reviewed considering phenotype quantification, genetic variant classification and functional assays of Merlin and its downstream pathways, studied by western blot in patient’s primary fibroblast. Principal component analysis and regression models were considered to evaluate the differences between severity and *NF2* germline mutations.

**Results:** The GSS was validated in the Spanish NF2 cohort. However, for some patients harboring mutations associated to mild and moderate phenotypes, GSS did not perform as well in predicting clinical outcomes as for pathogenic variants associated to severe phenotypes. We studied the possibility of modifying the mutation classification in GSS adding functional assays to evaluate the impact of pathogenic mutations on Merlin’s function. This revision help reducing variability within *NF2* mutation classes and moderately enhances the correlation between patients’ phenotype and the different prognosis parameters analyzed.

**Conclusions:** We validated the UK NF2 GSS in a Spanish NF2 cohort although a significant phenotypic variability was identified. The revision of the GSS, named FGSS, could be an added value to the classification of mosaic patients and patients showing mild and moderate phenotypes.

## 1. Introduction

Neurofibromatosis type 2 (NF2) (MIM 101000) is an autosomal-dominant syndrome caused by mutations in the *NF2* gene that affects 1 in 33,000-40,000 births worldwide. Approximately 50% of the cases are due to *de novo* mutations, while the other 50% are familial cases. NF2 typically presents with vestibular schwannomas (VS) in addition to multiples schwannomas in other cranial, peripheral and spinal nerves, as well as meningiomas, ependymomas and congenital cataracts accompanied by focal neurological deficits. Due to the development of bilateral VS, NF2 disease will progress in most patients causing hearing loss, tinnitus and balance problems[1]. Despite the benign nature of these tumors, their multiplicity and anatomical location entails high morbidity and early death. Therapeutic management is challenging in these patients due to the recurrence of treated tumors. Surgery, radiotherapy and Bevacizumab are the current gold standard treatments[2,3].

The clinical expression of the disease is highly variable, which makes the clinical management of these patients complex and challenging[4]. Phenotype differences found among patients already led to a clinical classification in which two subgroups of adult patients were established into Gardner and Wishart subtypes (mild and severe phenotypes, respectively). Further studies have resulted in the identification of several prognostic markers[5,6] and a good genotype-phenotype correlation[7–9].

In this context, the UK NF2 Reference Group established a Genetic Severity Score (GSS) to predict the severity of the disease based on the type of patient’s *NF2* germline variant[10]. This score stratifies NF2 patients in four groups: in groups 1A and 1B no pathogenic variant is identified in blood, and patients show a very mild phenotype, group 2 harbor splice-site mutations, large deletions, small in-frame in/dels, and missense mutations or truncating mutations in mosaicism, exhibiting mild (2A) and moderate (2B) phenotypes respectively. Group 3 carry truncating mutations in exons 2-13 of the *NF2* gene and present a severe phenotype. Therefore, the GSS represents a tool by which it is possible to establish a trend in NF2 prognosis from patient’s germline mutation and thus, improve the clinical follow-up and the genetic counselling offered to affected families[10,11].

Around 50% of NF2 *de novo* cases are mosaic (groups 1A and 1B) and present highly variable phenotypes[12,13]. Specifically, more than 15% of the *de novo* NF2 patients show bilateral VS before the age of 20 and with no mutation identified in blood[12,14]. Therefore, although the good performance of the UK NF2 GSS, this complicates the day-to-day application of GSS in the clinic as some mosaic cases may develop multiple tumors at early ages[6,15–17]. Similarly, some patients harboring splicing and missense mutations could also show significant differences in their clinical manifestations[7,8,18–23]. Hence, the GSS is useful to classify NF2 patients in general terms but could have room for improvement in stratifying some patients.

The *NF2* encodes for the protein Merlin. Merlin can be considered a scaffold protein as indirectly links F-actin, transmembrane receptors and intracellular effectors to modulate receptor mediated signaling pathways and integrates extracellular signals to modulate morphology, motility, proliferation and survival[24–26]. Understanding the impact of the pathogenic *NF2* variant on Merlin’s stability and on the regulation of its associated signaling pathways could be a way of better accounting for variant pathogenicity. In this context, analyzing the status of small GTPases (Rac1 and Ras), mTOR, PI3K/Akt and Hippo pathways in NF2 patients could help in determining the functional impact of the germline variant and therefore, this data could be used to improve the NF2 patient’s prognosis prediction capacity.

In this study, we present the validation of the UK NF2 GSS[10] in the NF2 patients attended in the Spanish National Reference Centre (CSUR) of Phacomatoses and we propose a revised version, called Functional Genetic Severity Score (FGSS), considering data obtained from functional assays and the predicted mutational effect on Merlin. We describe here the performance of the FGSS in comparison to GSS and show the changes observed in the behavior of some clinical prognosis markers analyzed in our cohort.

## 2. Materials and methods

All procedures performed were in accordance with the ethical standards of the IGTP Institutional Review Board, who approved this study and with the 1964 Helsinki declaration and its later amendments.

### Patients

Written informed consent was obtained from all individuals included in the study. Clinical data was recorded, five domains were assessed: patient demographics, tumor load, ocular features, hearing capacity and major interventions. Genetic test was performed using the customized I2HCP panel[27] in blood or tissue when available.

### Functional assay

Merlin’s downstream pathways were analyzed in 27 out of 52 NF2 patients through a Western Blot assay. Cultured fibroblasts from skin biopsies were processed as described in [28]. Three primary cultures from healthy donors were included as control samples (Ctrl1, Ctrl2 and Ctrl3) and grouped together with the tissue mosaic patients for the statistical analysis. Primary antibodies (Supplementary materials and methods) were incubated at 4°C overnight and secondary antibodies during 1h at room temperature (IRDye 680LT and IRDye 800CW, 1:1000 dilution, LI-COR). The statistical threshold that allowed the differentiation between controls or mosaic patients and the patients in group 3 (severe) was established through the ±2SD limits method.

### Statistics

All statistical analyses of the validation of the GSS were reproduced from the study of Halliday et al [10]. In order to study differences between protein expression levels in the functional assay, Kruskal-Wallis and Mann-Whitney U statistical tests were performed among genetic severity groups and pathogenic variant classes. Analysis of variance (ANOVA) of FGSS and *NF2* mutations was performed to test differences of severity between NF2 mutations groups. Backward stepwise regression analysis was performed to evaluate the contribution of Merlin and pERK levels in the ANOVA model. Spearman Correlation (ρ) was used to calculate the correlation between Merlin and pMerlin. Statistical analyses were performed using R software.

For more information, see Supplementary Materials and Methods.

## 3. Results

### 3.1. Validation of the UK NF2 Genetic Severity Score (GSS) in a Spanish cohort

Fifty-two patients from the Spanish National Reference Centre (CSUR) for Phacomatoses cohort were included in the GSS validation group. This cohort consisted of 19 men and 33 women, all of them with clinical diagnosis of NF2[29] at a mean age of 28.9 (±14.99), 44 patients showed bilateral VS, 39 presented spinal tumors, and 30 had intracranial meningiomas with a mean age of the cohort of 41,87 (±16.14). We stratified the whole cohort following the GSS classification. Nineteen cases of the cohort (34.6%) were assigned to group 1, including 6 confirmed tissue mosaics (group 1B, two or more affected tissue samples studied) and 13 to group 1A as presumed mosaics, since no mutation was identified in blood and affected tissues were not available. From the 33 patients with a constitutional *NF2* pathogenic variant, 9 patients were classified as 2A, 10 cases as 2B and 14 patients in group 3, with predicted phenotypes as mild, moderate and severe, respectively (Table 1-2).

**Table 1.**
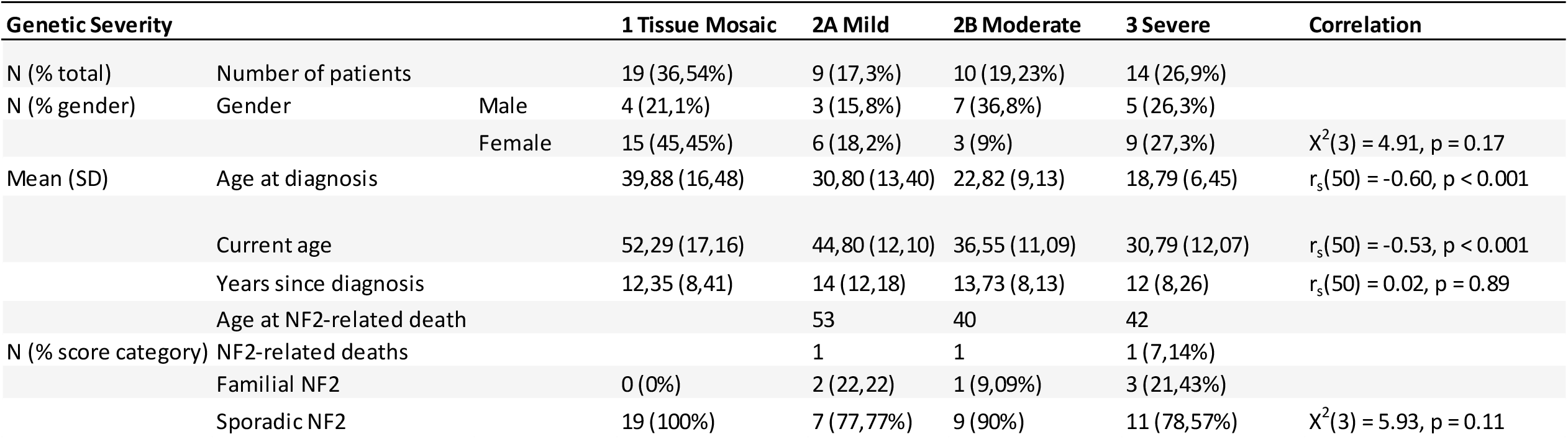
Demographic data according to Genetic Severity Score.

**Table 2.** Tumor burden, presence of ocular features and hearing outcome according to Genetic Severity Score (*Only patients from which data was available were taken into account in the analysis).

| Genetic Severity |  |  |  | 1 Tissue Mosaic | 2A Mild | 2B Moderate | 3 Severe | Statistics |
| --- | --- | --- | --- | --- | --- | --- | --- | --- |
| Number of patients |  |  |  | 19 | 9 | 10 | 14 |  |
| Tumor load | N (%) | Bilateral VS | | 11 (57,89%) | 9 (100%) | 10 (100%) | 14 (100%) | $\chi^2(1) = 8.38, p = 0.003$ |
| | | Unilateral VS | | 6 (31,58%) | 0 (0%) | 0 (0%) | 0 (0%) | $\chi^2(1) = 5.18, p = 0.022$ |
| | | Intracranial meningioma | | 11 (57,89%) | 2 (22,2%) | 7 (70%) | 10 (71,4%) | $\chi^2(1) = 0.79, p = 0.37$ |
| | | Intracranial Schwannoma | | 5 (26,3%) | 3 (33,3%) | 4 (40%) | 7 (50%) | $\chi^2(1) = 1.56, p = 0.21$ |
| | | Spinal meningioma | | 6 (31,58%) | 0 (0%) | 4 (40%) | 5 (35,7%) | $\chi^2(1) = 0.23, p = 0.62$ |
| | | Spinal Schwannoma | | 10 (52,63%) | 6 (66,7%) | 6 (60%) | 11 (78,6%) | $\chi^2(1) = 2.46, p = 0.11$ |
| | | Spinal ependymoma | | 4 (21,05%) | 1 (11,1%) | 4 (40%) | 5 (35,7%) | $\chi^2(1) = 1.23, p = 0.26$ |
| Ocular Features | N (%) | Epiretinal membranes |  | 0 (0%) | 0 (0%) | 0 (0%) | 0 (0%) | NA |
| | | Cataract | | 4 (21,05%) | 2 (22,2%) | 2 (22,2%) | 3 (21,42%) | $\chi^2(1) = 0.01, p = 0.90$ |
| | | Combined hamartoma | | 0 (0%) | 0 (0%) | 0 (0%) | 2 (14,28%) | $\chi^2(1) = 3.35, p = 0.06$ |
| | | Optic nerve meningioma | | 0 (0%) | 0 (0%) | 1 (10%) | 0 (0%) | $\chi^2(1) = 0.24, p = 0.62$ |
| | Mean (SD) | Total eye features | | 0,27 | 0,2 | 0,33 | 0,4 | $r_s(49) = 0.05, p = 0.69$ |
| Hearing Outcomes | N (%) | Hearing grade | 1 | 13 (68,4%) | 5 (55,5%) | 4 (40%) | 6 (42,9%) | $\chi^2(1) = 2.05, p = 0.15$ |
| | | | 2 | 1 (5,26%) | 0 (0%) | 0 (0%) | 0 (0%) | $\chi^2(1) = 1.33, p = 0.24$ |
| | | | 3 o 4 | 0 (0%) | 0 (0%) | 2 (20%) | 1 (7,1%) | $\chi^2(1) = 1.83, p = 0.17$ |
| | | | 5 | 2 (10,52%) | 2 (22,2%) | 1 (10%) | 4 (28,6%) | $\chi^2(1) = 1.01, p = 0.31$ |
| | | | 6 | 2 (10,52%) | 3 (33,3%) | 3 (30%) | 3 (21,4%) | $\chi^2(1) = 0.50, p = 0.47$ |
| | Mean (SD) | Age of loss of useful hearing | | 48,59 (18,57) | 38 (11,96) | 30,27 (9,88) | 26,23 (12,58) | $r_s(48) = -0.56, p < 0.001$ |
| Cutaneous manifestations | N (%) | | | 6 (31,57%) | 2 (22,2%) | 6 (60%) | 9 (64,3%) | $\chi^2(1) = 3,94, p = 0.047$ |

\* For group 2A and 3, only 9 out 10 and 11 out of 14 patients, respectively, have spinal magnetic resonance data

When analyzing demographic data (Table 1), we observed the mean age at diagnosis in individuals in group 1 (39.88 ±16.48) was statistically significantly different from patients of group 3 (18.79 ±6.45, p<0.001). The mean age at diagnosis for the group 2A was 30.80 (±13.40) years and slightly lower in group 2B (22.82 ±9.13) with no significant differences between these groups. Concerning the age at hearing loss, significant differences were also found between groups 1 and 3 (p<0.0005), observing a linear correlation between the age at hearing loss and the severity group established by the GSS (Table 2). Therefore, these results showed the same tendency as the English cohort and that features related to poor prognosis such as the age at diagnosis and age at hearing loss correlated with the four groups established.

When studying tumor burden, the highest incidence of intracranial meningiomas was found in groups 2B and 3 (∼70%), being lower in group 2A (22.2%) and, unexpectedly, considerably high in group 1 (57.9%). Similarly, intracranial schwannomas were also represented in patients classified in groups associated to mild phenotypes. Spinal meningioma was reported in groups 1, 2B and 3 of our cohort, specifically, six mosaic patients of our cohort developed multiple spinal meningiomas. A fairly even distribution of the presence of spinal schwannomas was seen in the groups 1 and 2 (52 to 66%) and a higher incidence was found in group 3 (78.6%) with not statistically significant differences. Regarding spinal ependymomas, more incidence was found in patients from groups 2B and 3 (40% and 35.7% respectively), but also a considerable variability in the other groups and no clear linear trend was observed. Finally, considering the number of major interventions in our cohort we did not observe significant differences between groups (Table 2, Supplementary Table 1-2).

### 3.2. Functional analysis of Merlin and associated signaling pathways in patient’s skin fibroblasts

A functional assay in patient’s primary cultured fibroblasts was developed in order to analyze the activation state of Merlin and some of its downstream pathways to better interpret the pathogenicity of *NF2* constitutional variants in relation to the phenotype variability observed within GSS categories. In particular, we analyzed PAK1 and RAC total protein amount, and total protein and their phosphorylated forms of Merlin, ERK, PKA, YAP and AKT.

As expected, all patients that harbored a germline pathogenic variant and therefore, with one *NF2* mutated allele, showed lower Merlin levels compared to healthy controls or tissue mosaics (p<0.05). No significant differences were found among groups with a pathogenic variant in the *NF2* gene, regardless the variants’ type or location within the gene, meaning that the effect on Merlin levels were very similar in patients carrying truncating and splicing variants or large deletions (Figure 1A and 1B, Supplementary Table 1). When studying the status of phosphorylated Merlin (pMerlin), we observed the same tendency as Merlin, suggesting that decreased levels of pMerlin could be due to the lower levels of Merlin rather than a dysregulation of Merlin activation (Supplementary Figure 1).

**Figure 1.**
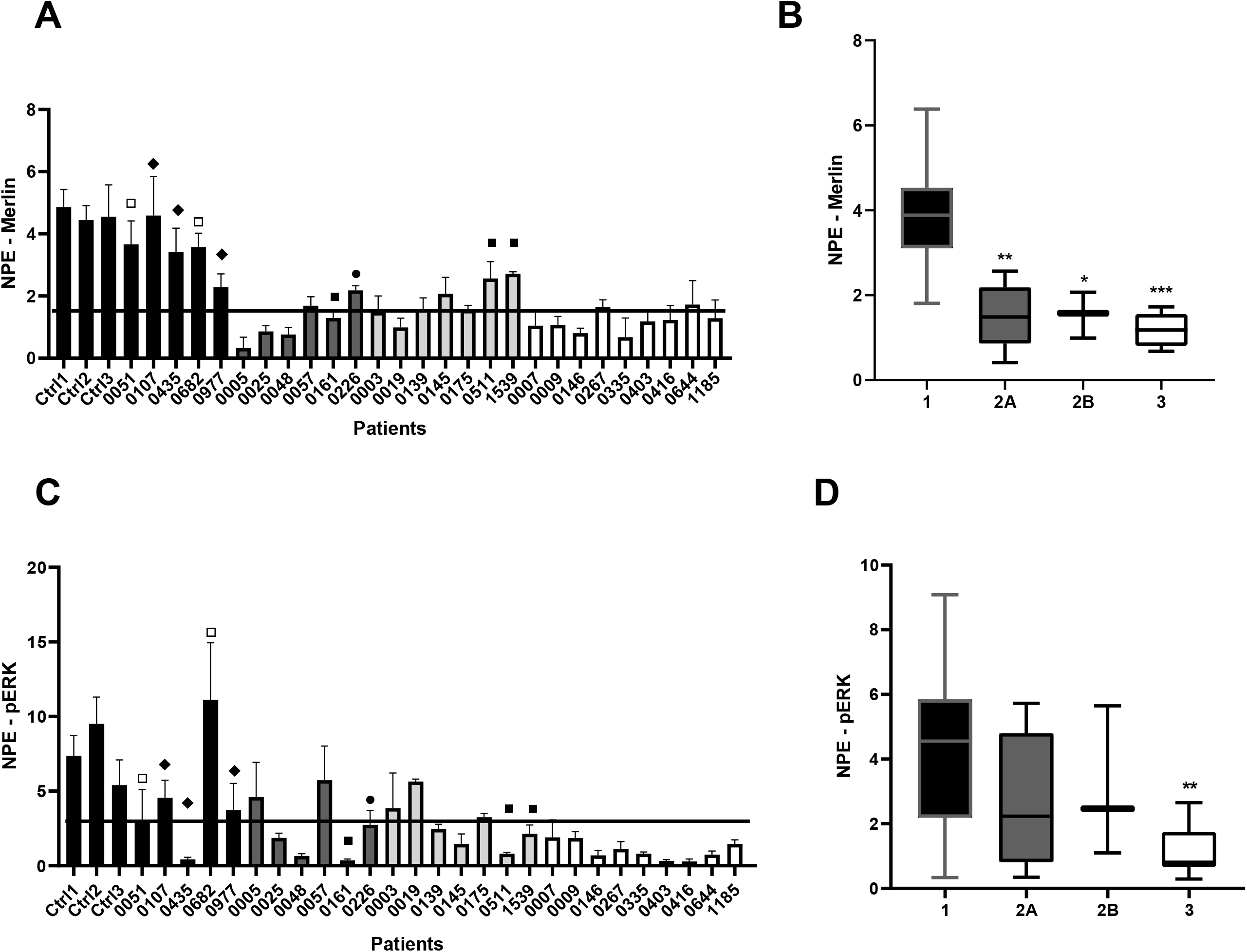
Merlin and pERK levels in patient’s fibroblasts according to the GSS. **A)** Merlin levels in NF2 patients’ fibroblasts. **1B)** Average of Merlin levels according to Genetic Severity Groups. **1C)** pERK levels in NF2 patients’ fibroblasts; **1D)** Average of pERK levels according to Genetic Severity Groups. Column’s grey scale indicates the Genetic Severity Group; NPE stands for Normalized Protein Expression; Bars represent the SD from three independent experiments. (*p <0.05; **p <0.005; ***p <0.001). Symbol legend: ▢ presumed mosaicism; ▪ generalized mosaicism; ♦ tissue mosaicism; ᴑ missense.

Results did not indicate any differences in the status of the PI3K, PAK, YAP or RAC pathways, neither in the total protein levels nor in the phosphorylated forms. In contrast, pERK levels in patients’ fibroblasts with truncating mutations were statistically significantly lower than pERK levels in tissue mosaics or healthy controls), while ERK levels remained constant between groups (Figure 1C-D, Supplementary Figure 2). These results allowed a statistically significant discrimination of patients with severe phenotypes from healthy controls (p=0.0042). The group with intermediate phenotype showed greater variability, as did the group of mosaic patients (SD 2A=1.999, SD 2B=2.332, 1A&B=2.624).

### 3.3. Reviewing GSS mutation categorization

Due to the unexpected tendency among groups 1 and 2 in the clinical manifestations of the studied cohort we revised the GSS considering the predicted mutation effect on Merlin, its position within the gene, and its effect on Merlin and pERK activity based on functional assays results.

Firstly, qualitative clinical data recorded was coded to quantitative variables in order to measure the NF2 phenotype independently of the *NF2* mutation type and the mosaicism status. Quantification of the phenotype was based on well-known NF2 prognosis markers (Supplementary Table 3). The NF2 phenotype was presented in a numeric ten-scale based on the sum of the transformed quantitative variables. In this way, a patient younger than 25 years old, presenting bilateral VS, peripheral schwannomas, and multiple spinal/cerebral injuries was classified with the most severe score (10). Secondly, we analyzed the relation between the GSS predicted phenotype and the outcome of phenotype quantification. We observed that some tissue mosaic patients did not show the expected very mild phenotype, patients carrying large deletions including or excluding the 5’ of the gene did not show phenotypic differences, and neither did the patients harboring splice variants in exons 1-7 or 8-13 (Supplementary Table 4).

Taking into account the mentioned observations, we revised GSS categorization of *NF2* mutations and propose a modified criteria rated in six mutation classes, similarly to GSS subcategories (1A/B, 2A/B and 3), and referred as Functional Genetic Severity Score (FGSS). In general terms, presumed mosaics or patients carrying a Ring22 were scored as 1 and related to very mild phenotypes, while truncating mutations in exons 2-13 were associated to a severe phenotype and scored as 6. Other types of pathogenic variants were scored in between considering if the variant induced a frameshift alteration and the proportion of cells carrying the mutation. However, we did not take into account the position of the altered exons, except for exon 1, since no differences were observed in Merlin levels when comparing mutations affecting 5’ or 3’ of the *NF2* gene (Table 3, Supplementary Table 4, Supplementary Materials & Methods).

**Table 3.**
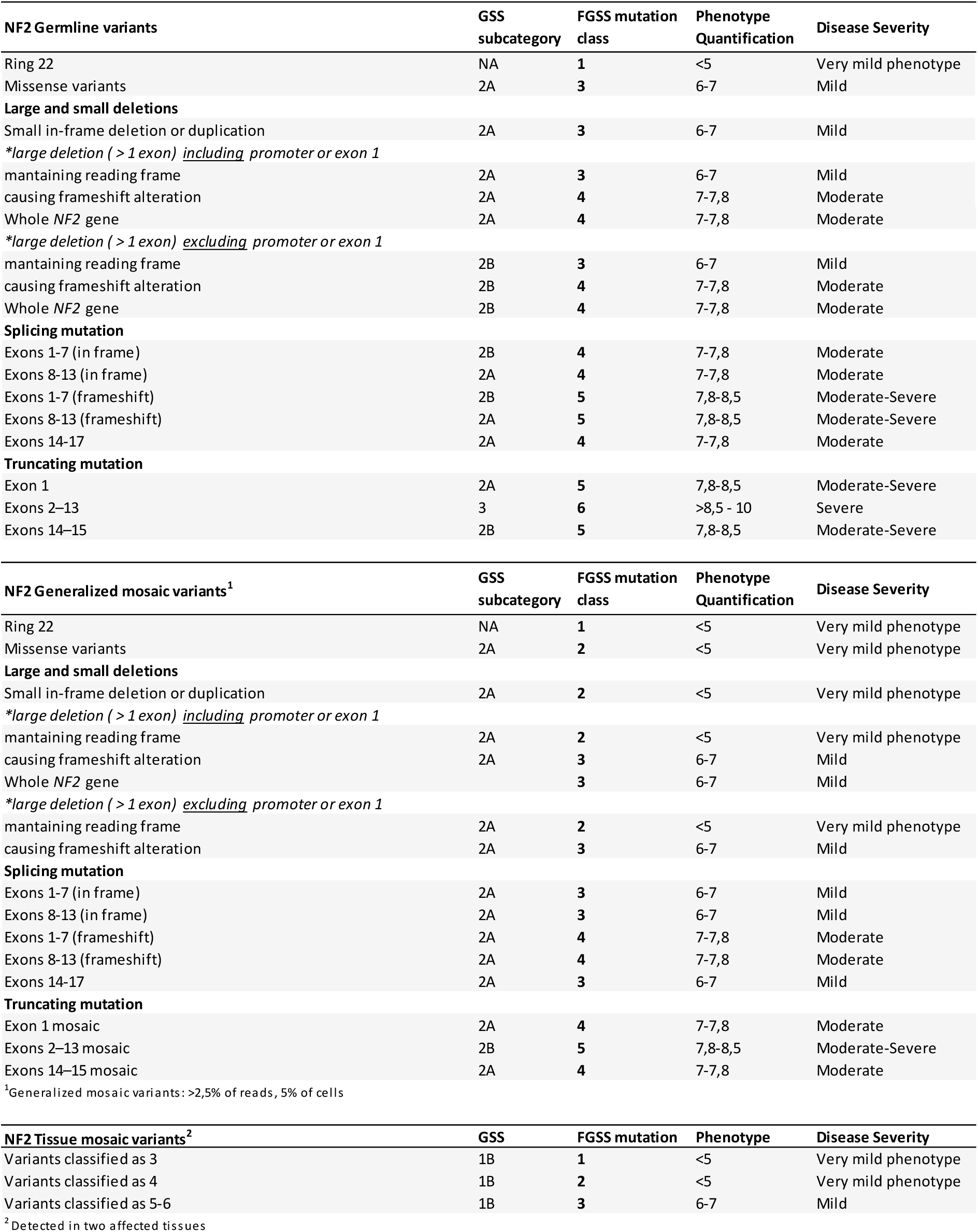
Categorization of NF2 mutation according to Functional Genetic Severity Score.

| NF2 Germline variants | GSS subcategory | FGSS mutation class | Phenotype Quantification | Disease Severity |
| --- | --- | --- | --- | --- |
| Ring 22 | NA | 1 | <5 | Very mild phenotype |
| Missense variants | 2A | 3 | 6-7 | Mild |
| **Large and small deletions** |  |  |  |  |
| Small in-frame deletion or duplication | 2A | 3 | 6-7 | Mild |
| *\*large deletion (> 1 exon) including promoter or exon 1* |  |  |  |  |
| maintaining reading frame | 2A | 3 | 6-7 | Mild |
| causing frameshift alteration | 2A | 4 | 7-7,8 | Moderate |
| Whole NF2 gene | 2A | 4 | 7-7,8 | Moderate |
| *\*large deletion (> 1 exon) excluding promoter or exon 1* |  |  |  |  |
| maintaining reading frame | 2B | 3 | 6-7 | Mild |
| causing frameshift alteration | 2B | 4 | 7-7,8 | Moderate |
| Whole NF2 gene | 2B | 4 | 7-7,8 | Moderate |
| **Splicing mutation** |  |  |  |  |
| Exons 1-7 (in frame) | 2B | 4 | 7-7,8 | Moderate |
| Exons 8-13 (in frame) | 2A | 4 | 7-7,8 | Moderate |
| Exons 1-7 (frameshift) | 2B | 5 | 7,8-8,5 | Moderate-Severe |
| Exons 8-13 (frameshift) | 2A | 5 | 7,8-8,5 | Moderate-Severe |
| Exons 14-17 | 2A | 4 | 7-7,8 | Moderate |
| **Truncating mutation** |  |  |  |  |
| Exon 1 | 2A | 5 | 7,8-8,5 | Moderate-Severe |
| Exons 2-13 | 3 | 6 | >8,5 - 10 | Severe |
| Exons 14-15 | 2B | 5 | 7,8-8,5 | Moderate-Severe |

| NF2 Generalized mosaic variants<sup>1</sup> | GSS subcategory | FGSS mutation class | Phenotype Quantification | Disease Severity |
| --- | --- | --- | --- | --- |
| Ring 22 | NA | 1 | <5 | Very mild phenotype |
| Missense variants | 2A | 2 | <5 | Very mild phenotype |
| **Large and small deletions** |  |  |  |  |
| Small in-frame deletion or duplication | 2A | 2 | <5 | Very mild phenotype |
| *\*large deletion (> 1 exon) including promoter or exon 1* |  |  |  |  |
| maintaining reading frame | 2A | 2 | <5 | Very mild phenotype |
| causing frameshift alteration | 2A | 3 | 6-7 | Mild |
| Whole NF2 gene |  | 3 | 6-7 | Mild |
| *\*large deletion (> 1 exon) excluding promoter or exon 1* |  |  |  |  |
| maintaining reading frame | 2A | 2 | <5 | Very mild phenotype |
| causing frameshift alteration | 2A | 3 | 6-7 | Mild |
| **Splicing mutation** |  |  |  |  |
| Exons 1-7 (in frame) | 2A | 3 | 6-7 | Mild |
| Exons 8-13 (in frame) | 2A | 3 | 6-7 | Mild |
| Exons 1-7 (frameshift) | 2A | 4 | 7-7,8 | Moderate |
| Exons 8-13 (frameshift) | 2A | 4 | 7-7,8 | Moderate |
| Exons 14-17 | 2A | 3 | 6-7 | Mild |
| **Truncating mutation** |  |  |  |  |
| Exon 1 mosaic | 2A | 4 | 7-7,8 | Moderate |
| Exons 2-13 mosaic | 2B | 5 | 7,8-8,5 | Moderate-Severe |
| Exons 14-15 mosaic | 2A | 4 | 7-7,8 | Moderate |

| NF2 Tissue mosaic variants<sup>2</sup> | GSS | FGSS mutation | Phenotype | Disease Severity |
| --- | --- | --- | --- | --- |
| Variants classified as 3 | 1B | 1 | <5 | Very mild phenotype |
| Variants classified as 4 | 1B | 2 | <5 | Very mild phenotype |
| Variants classified as 5-6 | 1B | 3 | 6-7 | Mild |

<sup>1</sup>Generalized mosaic variants: >2,5% of reads, 5% of cells<sup>2</sup>Detected in two affected tissues

### 3.4. Analysis of the added value of incorporating functional information to GSS

When stratifying our cohort with the proposed criteria, patients harboring genetic variants scored as 5 or 6 showed a severe phenotype, while patients harboring *NF2* mutation class 4 behaved fairly similar, although less extravestibular lesions were observed. In contrast, patients classified as class 3 presented a greater clinical heterogeneity since it included generalized and tissue mosaicisms as well as patients carrying constitutional variants associated to a mild phenotype. Finally, patients of classes 1 or 2 showed very mild phenotypes since these groups considered presumed mosaic and tissue mosaic patients harboring less deleterious variants. The FGSS showed light stronger correlation among classes when studying the appearance of vestibular schwannomas, the presence of intracranial meningiomas, as well as in hearing outcomes and cutaneous manifestations. The age at first surgery and several parameters analyzed related to major interventions also showed larger means between classes when compared to the GSS (Table 2 and Supplementary Table 1,5-7). In addition, the proposed revision to classify patient’s phenotype based on the *NF2* genetic variant reduced the intragroup variability, improved mosaic patient’s classification and moderately improved the correlation between patients’ phenotype and some prognosis parameters analyzed (Figure 2, Supplementary Table 5-8). All these phenomena could indicate that the revised criteria might be an added value to the GSS.

**Figure 2.**
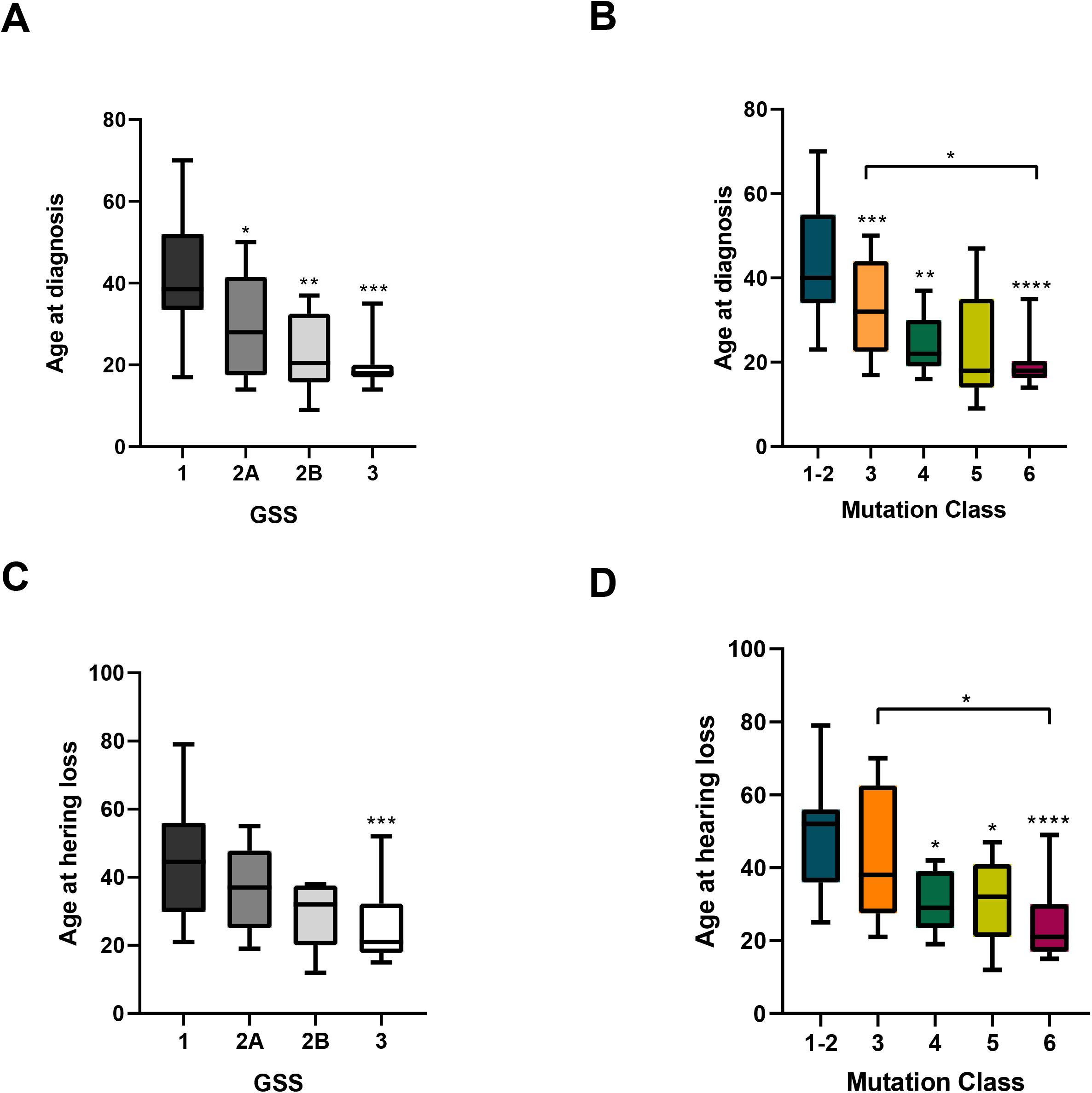
Comparison of the age diagnosis and age at hearing loss according to GSS or FGSS. **A)** Average age at diagnosis according to the GSS classification. **B)** Average age at diagnosis according to the FGSS. **C)** Average age at hearing loss according to the GSS; **D)** Average age at hearing loss according to the FGSS. Column’s grey scale indicates the Genetic Severity Group; Blue stands for the healthy controls and mutation classes 1 and 2; orange stands for mutation class 3; dark green stands for mutation class 4; light green stands for mutation class 5 and purple stands for mutation class 6. Bars represent the SD; (*p<0.05; **p<0.005; ***p<0.001, ****p<0.0005).

Similarly, Merlin and pERK levels showed the same trend as when classified through the GSS but the intragroup variability decreased in classes associated to mild phenotypes. In addition, Merlin levels were similar in groups 4-6 and significantly lower than groups 1-3 (p<0.005), since these three last groups contained mosaic patients. Furthermore, the levels of pERK in class 5 and 6 and were statistically significantly lower compared to class 4 and below (p<0.005). As mentioned before, class 3 showed higher variability due to the phenotypic variability detected in tissue mosaic patients (σ=1.884) (Figure 3, Supplementary Table 8).

**Figure 3.**
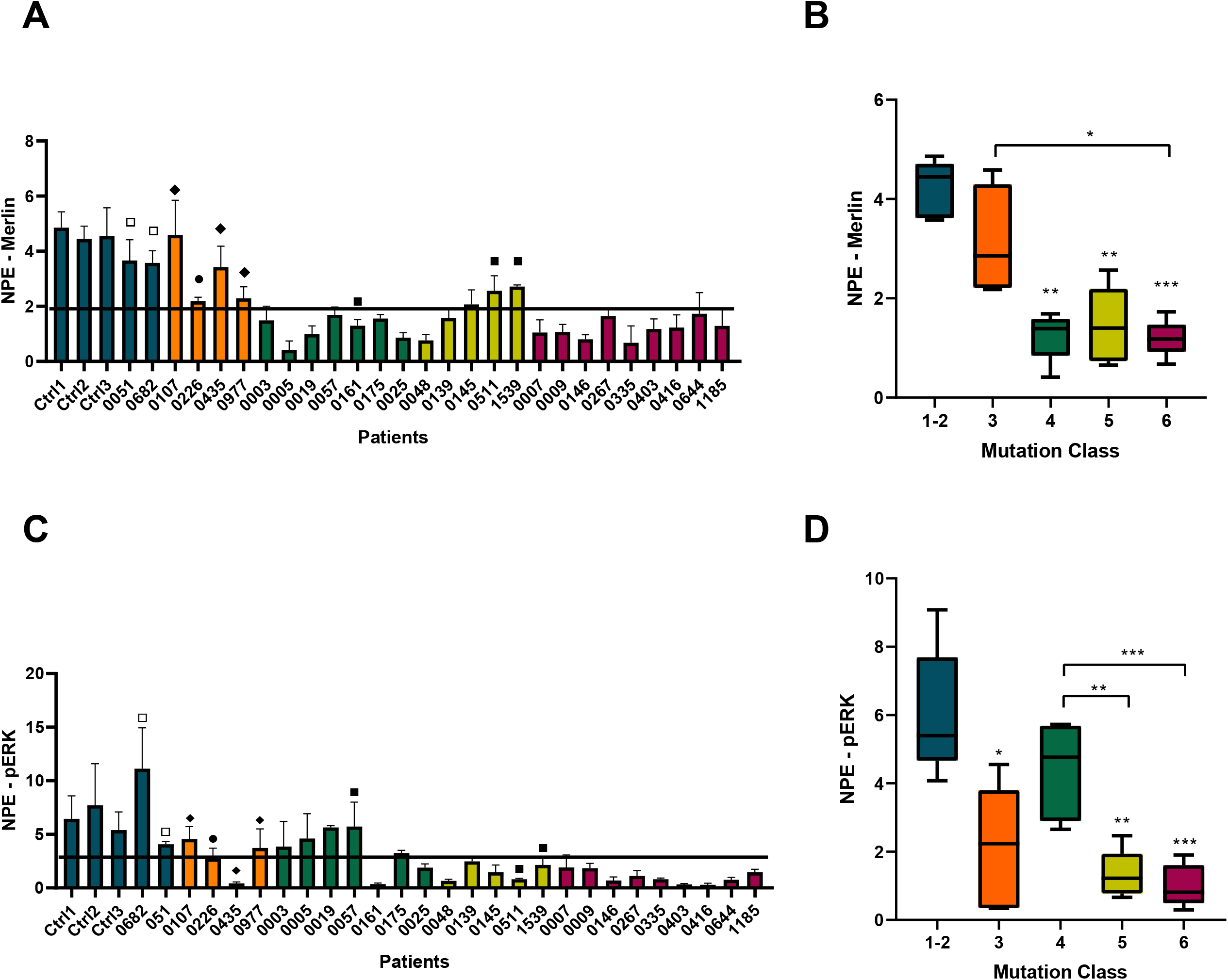
Merlin and pERK levels in patient’s fibroblasts according to the FGSS. **A)** Merlin levels in NF2 patients’ fibroblasts. **B)** Average of Merlin levels according to Mutation Class. **C)** pERK levels in NF2 patients’ fibroblasts; **D)** Average of pERK levels according to Mutation Class. Blue stands for the healthy controls and mutation classes 1 and 2; orange stands for mutation class 3; dark green stands for mutation class 4; light green stands for mutation class 5 and purple stands for mutation class 6. NPE stands for Normalized Protein Expression; Bars represent the SD from three independent experiments; (*p<0.05; **p<0.005; ***p<0.001). Symbol legend: ▢ presumed mosaicism; ▪ generalized mosaicism; ♦ tissue mosaicism; ᴑ missense.

In addition, we explored the variability of phenotypical data by principal component analysis (PCA) (Supplementary Figure 3). The PCA-biplot suggested that patients in the first quadrant mainly had *NF2* mutations of classes 4, 5 and 6, early age at diagnosis, presented extravestibular affection, vestibular and peripheral schwannomas, whereas patients with negative values in the PCA-biplot presented late age at diagnosis and mainly NF2 mutations of class 1. To evaluate the tendency observed using the FGSS and considering the different number of genetic subcategories between GSS and FGSS, we performed a regression model (ANOVA) between the quantitative phenotype and *NF2* disease-causing variant class to evaluate quantitatively the NF2 phenotype in base of the *NF2* variant (Supplementary Table 9A, Shapiro-Wilk p=0.67, Levene test p=0.16). We observed that NF2 phenotype could be explained partially by the *NF2* germline variant, specifically by mutations classified as 3 or higher scores (R2=0.38, p=0.0001). Therefore, mutations classified as 6 were associated to patients showing the severest phenotypes (phenotype>8.5), while mutations classified as 3 were associated to patients presenting milder phenotypes (phenotype ∼7). These results agreed with the observations that some mosaic patients showed a severe phenotype. In this regard, the fact that the *NF2* pathogenic variant could explain no more than 40% of the phenotype, indicated that many other variables could explain the resulting phenotype of the patient. Due to the differences observed in the ERK activation, the incorporation of functional data from 27 patients to the statistical model was analyzed. Yet, using this sample set, the incorporation of pERK levels into the ANOVA model, in addition to *NF2* germline mutation class data, seemed not to improve the capacity to explain the NF2 phenotype (p>0.05, Supplementary Table 9).

In contrast, the regression model between GSS and *NF2* disease-causing variant showed a R2=0.32 (p=0.001) and was only statistically significant in groups 2B and 3 (Supplementary Table 9B). Unfortunately, none of these models could explain presumed mosaics or some of the tissue mosaic patients. These results indicated that FGSS would improve the contribution of the *NF2* variant in predicting the NF2 phenotype and could improve the NF2 patient’s prognosis prediction capacity in comparison to GSS.

## 4. Discussion

The clinical management of the NF2 is complex as not only requires a specialized multidisciplinary team, but also presents challenges due to the phenotypic variability found among patients. A huge effort has been done to determine NF2 prognosis markers[5,6] and to establish a good genotype-phenotype relationship in order to be able to predict NF2 prognosis[7–9]. Improvement in determining patient’s prognosis allows an optimization of the patients’ clinical follow-up and ameliorate the genetic counseling offered to affected families.

The UK NF2 Genetic Severity Score (GSS)[10] is an objective tool to predict the trend of NF2 prognosis based on the type of patient’s germline genetic variant. The application of the GSS in our Spanish cohort proved significant differences in several prognosis markers between groups 1 and 3, and thus, allowing the GSS validation to determine the prognosis of patients carrying mutations associated to severest phenotypes. However, the GSS has difficulties, within our cohort, to classify accurately enough patients harboring mutations associated to mild or moderate NF2 phenotypes, neither to differentiate mosaic patients between them, since not all of this group showed a mild clinical presentation. Certain inconsistencies have been detected, as the case of four patients from group 2A and one mosaic patient (group 1B), who were clinically diagnosed nearby the age of 20 and that have recently developed multiple central nervous system tumors in addition to vestibular schwannomas, three of them before the thirties. These disparities could be due to the cohort size (n=52) or due to a possible bias since the largest number of patients in the cohort belong to the severe phenotype group. In contrast to the UK health system, the Spanish Reference Center is responsible for the management of NF2 patients of its sanitary region, and patients attended in other centers are generally only referred when the clinical management is complex. Hence, our cohort could be biased due to the low representation in mild phenotypes. In addition, other factors need to be taken into consideration in addition to *NF2* pathogenic variant as there is a wide evidence in the mild-moderate phenotypes heterogeneity, even in patients harboring the same *NF2* variant[8,9,12,13,19,20,30], as observed also in our statistical model. The GSS, although being appropriate to establish general trends, there is scope for improvement with other sort of data to establish a personalized NF2 prognosis score.

Understanding the status of Merlin and its regulated pathways might provide data that could help in the interpretation of the NF2 genotype-phenotype relationship. With this aim, a functional assay in patient’s primary cultured fibroblast was developed. As expected, Merlin levels showed significant differences between group 1 (no mutation identified in blood), compared to the groups harboring a constitutional *NF2* genetic variant. Furthermore, no differences were found regardless the type or the location of mutation within the gene. Although evidences in Schwann cells or immortalized tumor cells show the opposite effect, in this study, significant lower levels of pERK in Merlin haploinsuficient primary fibroblasts in comparison with healthy controls and tissue mosaics were observed[25,31–33]. Hence, these results, if validated in other cohorts and in larger sample sizes, could allow to establish that pERK levels in patient’s fibroblast are associated to severe phenotype, and thus, used as a NF2 prognosis marker.

In this study, we decided to quantify the NF2 phenotype using a ten-scale system based on well-known NF2 prognosis markers[5]. This scale allows not only an easy quantification of the NF2 phenotype, presumably more accurate than using a qualitative system (mild-moderate-severe), but also the quantification of mosaics’ phenotype. In addition, this quantitative method enables the modelling of the NF2 phenotype according to *NF2* genetic variant, as has been done in other studies[34–36].

Subsequently, with the aim to improve the NF2 prognosis capacity based on the germline variant, and also to assess if functional data could be incorporated to the UK NF2 GSS, we reviewed the classification of variants proposed by Halliday et al[10]. Similarly to GSS, we propose to consider the mosaicism extension, the published NF2 genotype-phenotype evidences[7,8,10,12,37], in addition to the predicted mutation effect on Merlin function and the functional assays results. We classified *NF2* genetic variants in six classes, where truncating mutations in exons 2-13 were scored 6, missense and in frame deletions as 3, presumed mosaics or patients carrying a Ring22 as 1, while the other type of pathogenic variants were scored in between. The main dissimilarities to GSS are the classification of large deletions and splicing variants, were the position in *NF2* gene were contemplated differently, since several evidences indicate that splicing variants affecting the N-term or C-term of NF2 gene are associated to both severe and moderate phenotypes, respectively [5,7,8,20–22], and variants that do not alter the reading frame have lower scores in comparison as those generating a frameshift alteration, assuming that in frame alterations could generate an hypomorphic Merlin preventing the activation of nonsense-mediated mRNA decay.

Within our cohort, the reviewed GSS, called Functional Genetic Severity Score (FGSS) seems to reduce the intragroup variability, helps the mosaic patient’s stratification and slightly upgrades the correlation between patients’ phenotype and the different prognosis parameters analyzed in relation to the GSS. Moreover, the modeling of the NF2 phenotype in terms of the *NF2* mutation classification in our cohort corroborates that *NF2* variants can explain partially patient’s phenotype and thereby, the GSS with this added value might be more precise when determining a prognosis trend in NF2 patients.

Similarly to the GSS, FGSS is unable to classify properly presumed and some tissue mosaic patients. Although mosaic patients are generally associated to mild phenotypes, some of these patients show a rather severe phenotype, indicating that the extension of mosaicism should also be considered. Regarding this point, we observed that *NF2* analysis on blood may not represent the affection degree given that it is known that tissue mosaic NF2 patients are at 6% of risk of transmitting the disease[14], and, moreover, the genetic tests in this study show that in some patients the NF2 mutation was undetectable in blood but was present in the skin fibroblasts (Supplementary Figure 4). These outcomes could indicate that hematopoietic primordial stem cells harboring *NF2* mutations may have a selective growth disadvantage over normal stem cells, reducing their representation in this tissue, contrarily to what it has been shown for NF1 mutations [38,39].

In addition, Merlin and pERK levels behave similar using the GSS and the FGSS, but with the latter show an increased correlation along with the NF2 phenotype and a decreased intragroup variability in groups associated to mild phenotypes. However, although pERK levels could represent a potential NF2 prognosis marker, its incorporation to the genetic score do not improve the capacity to explain NF2 phenotype. This could be due to the small sample set included in the model (n=27) or because the contribution of pERK on NF2 phenotype is too low when compared to the contribution of the *NF2* variant in the phenotype description, thus, its addition does not increase the statistical significance of the model. Although discouraging, these results should be tested in larger sets of samples because of the promising tendency of pERK levels.

To conclude, with the data obtained from this study, we propose a review of the UK NF2 GSS to help establishing a personalized trend in NF2 prognosis. This revision considers not only the type of germline pathogenic variant, the extent of mosaicism, genotype-phenotype evidences and statistically significant correlations already published, but also the predicted mutation effect on Merlin’s function and its effect on Merlin and pERK activity.

## Supporting information

Supplementary Figure 1. pMerlin and Merlin levels in patient's fibroblasts. NPE stands for Normalized Protein Expression; Bars represent the SD from t

Supplementary Figure 3. PCA-biplot of phenotypical NF2 data by NF2-mutation groups. VS: Vestibular Schwannoma; Age Dx: Age at diagnosis; PS: Periphera

Supplementary Figure 4. Sanger sequencing comparison between blood and fibroblast samples of a NF2 patient.

Supplementary Table 1. NF2 Patients summary.

Supplementary Table 2. Major interventions in relation to Genetic Severity Score.

Supplementary Table 3. NF2 Phenotype quantification.

Supplementary Table 4. Revising pathogenicity associated to NF2 mutations according to GSS. Patients with an unexpected phenotype according to GSS are

Supplementary Table 5. Demographic data according to Functional Genetic Severity Score.

Supplementary Table 6. Tumor burden, presence of ocular features and hearing outcome according to Functional Genetic Severity Score

Supplementary Table 7. Major interventions in relation to Functional Genetic Severity Score.

Supplementary Table 8. Intragroup Variability of Merlin, pERK, age at diagnosis and age at hearing loss.

Supplementary Table 9. A) FGSS and NF2 disease-causing variant regression model. B) FGSS and NF2 disease-causing variant regression model.

## Data Availability

Clinical and genetic data from a NF2 Spanish cohort is available.

## ACKNOWLEDGEMENTS

We thank the HGTP Clinical Services and staff for their collaboration in generating and collecting patient’s clinical data. We would like to acknowledge the constant support of the different NF lay associations: Asociación de Afectados de Neurofibromatosis, Chromo22 and ACNefi.

The authors declare no conflicts of interests.

## Supplementary Material

**Supplementary Figure 1**. pMerlin and Merlin levels in patient’s fibroblasts. NPE stands for Normalized Protein Expression; Bars represent the SD from three independent experiments.

**Supplementary Figure 2**. Levels of proteins involved in the NF2 downstream pathway in patients’ fibroblasts. NPE stands for Normalized Protein Expression; Bars represent the SD from three independent experiments.

**Supplementary Figure 3**. PCA-biplot of phenotypical NF2 data by NF2-mutation groups. VS: Vestibular Schwannoma; Age Dx: Age at diagnosis; PS: Peripheral Schwannoma; EVA: Extra vestibular Affection. Dim1 (Dimension 1); Dim2 (Dimension 1)

**Supplementary Figure 4**. Sanger sequencing comparison between blood and fibroblast samples of a NF2 patient.

**Supplementary Table 1**. NF2 Patients summary.

**Supplementary Table 2**. Major interventions in relation to Genetic Severity Score.

**Supplementary Table 3**. NF2 Phenotype quantification.

**Supplementary Table 4**. Revising pathogenicity associated to *NF2* mutations according to GSS. Patients with an unexpected phenotype according to GSS are highlighted in bold in the Reasoning column.

**Supplementary Table 5**. Demographic data according to Functional Genetic Severity Score.

**Supplementary Table 6**. Tumor burden, presence of ocular features and hearing outcome according to Functional Genetic Severity Score.

**Supplementary Table 7**. Major interventions in relation to Functional Genetic Severity Score.

**Supplementary Table 8**. Intragroup Variability of Merlin, pERK, age at diagnosis and age at hearing loss.

**Supplementary Table 9**. A) FGSS and *NF2* disease-causing variant regression model. B) FGSS and *NF2* disease-causing variant regression model.

## Notes

### Competing Interest Statement

The authors have declared no competing interest.

### Funding Statement

This works has been funding by Spanish NF lay association through 'Fundacion Proyecto Neurofibromatosis', Spanish Fundacion FEDER for Rare Disease Investigation and by the Catalonia Goverment: AGAUR 2017 SGR 496

