## Supplementary figures and images for "Revisiting the UK Genetic Severity Score for NF2: a proposal for adding functional information"

### Supplemental Data 1

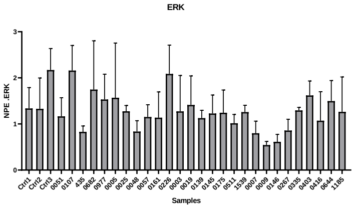

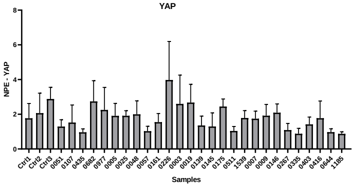

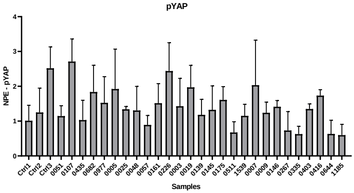

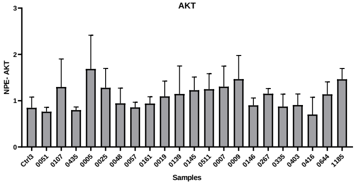

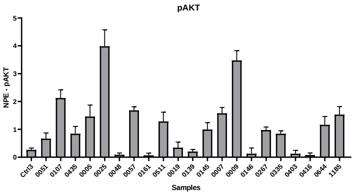

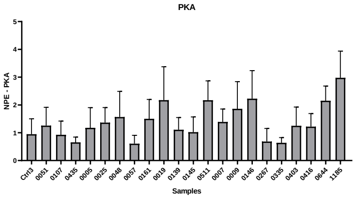

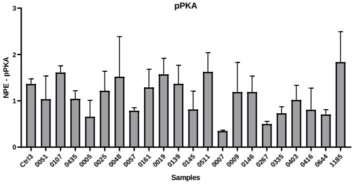

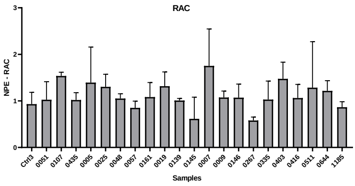

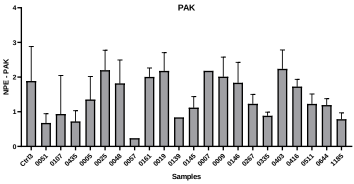

### Supplementary Figure 1. pMerlin and Merlin levels in patient's fibroblasts. NPE stands for Normalized Protein Expression; Bars represent the SD from t

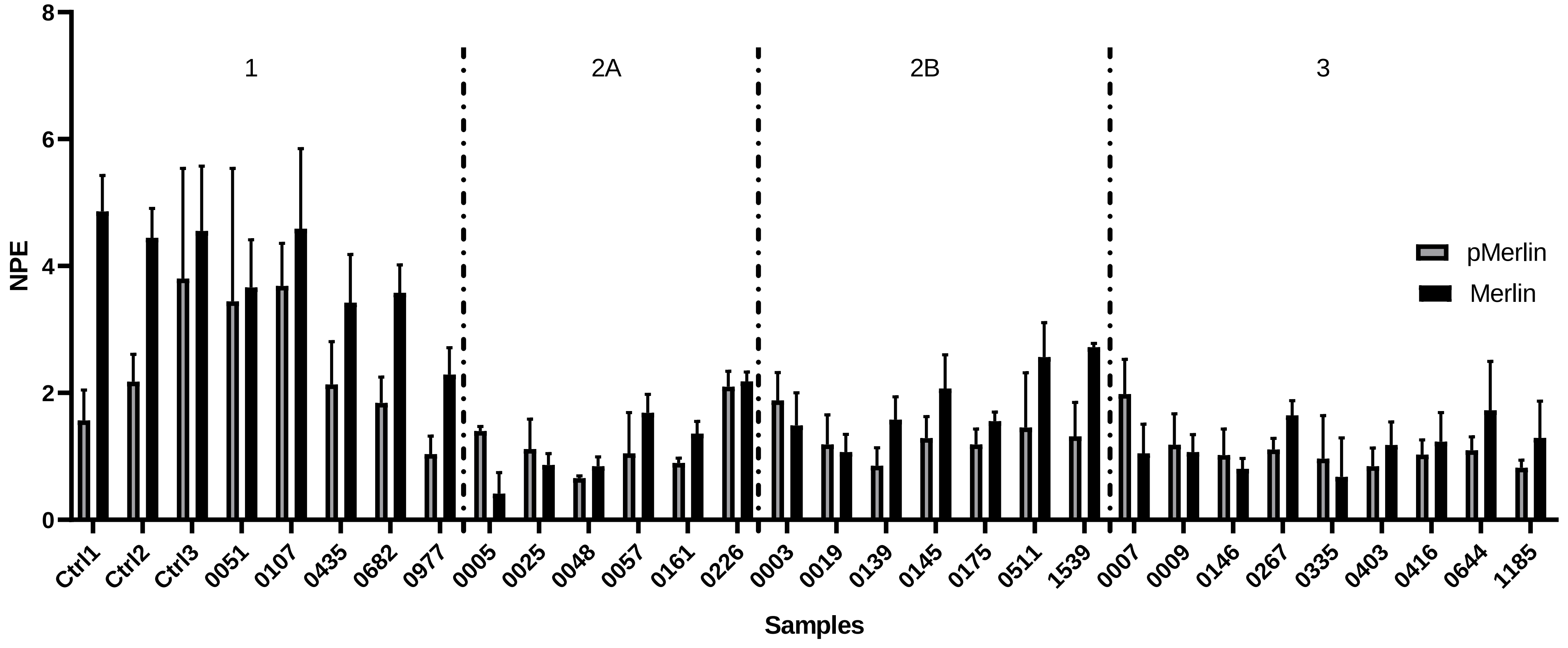

### Supplementary Figure 3. PCA-biplot of phenotypical NF2 data by NF2-mutation groups. VS: Vestibular Schwannoma; Age Dx: Age at diagnosis; PS: Periphera

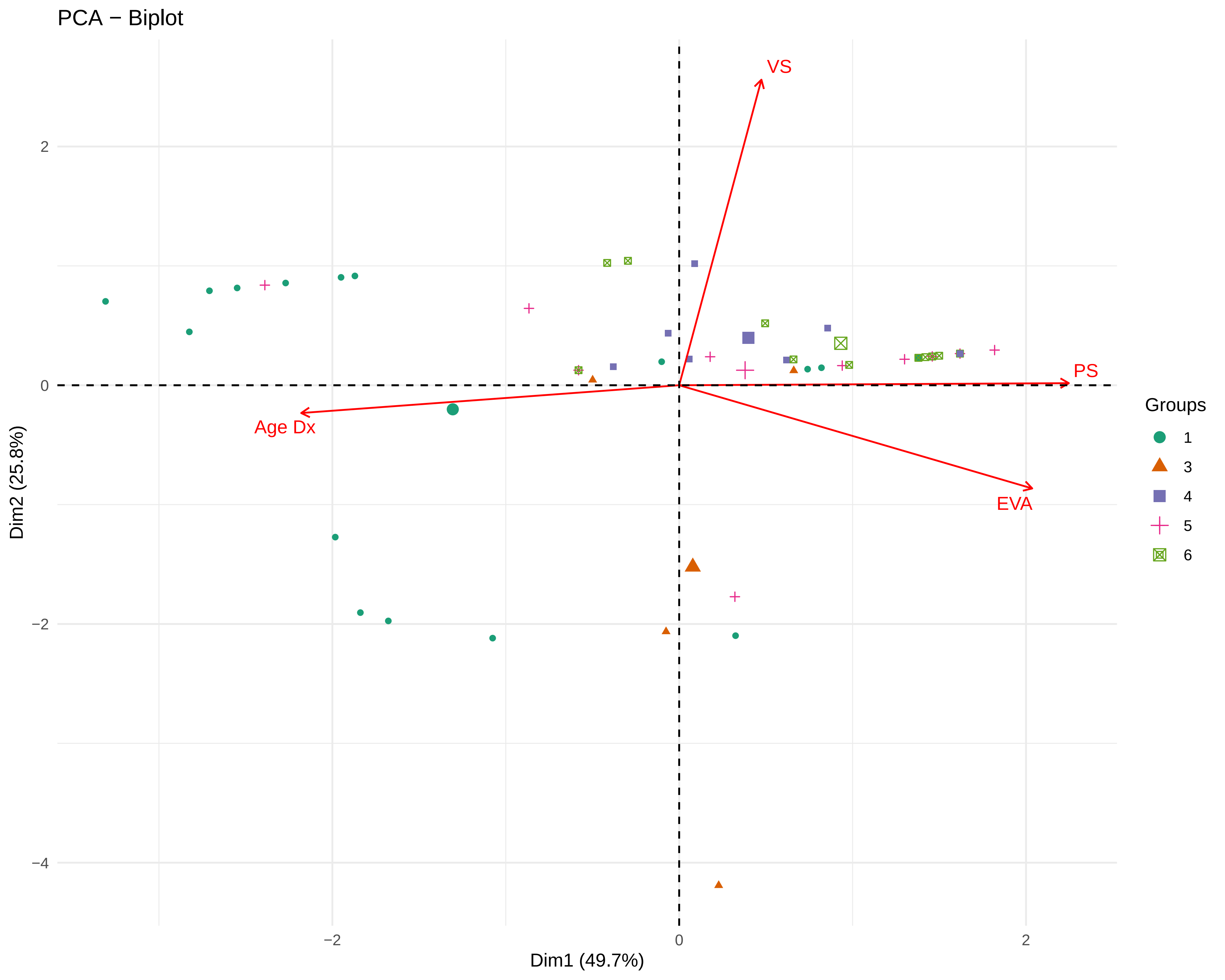

### Supplementary Figure 4. Sanger sequencing comparison between blood and fibroblast samples of a NF2 patient.

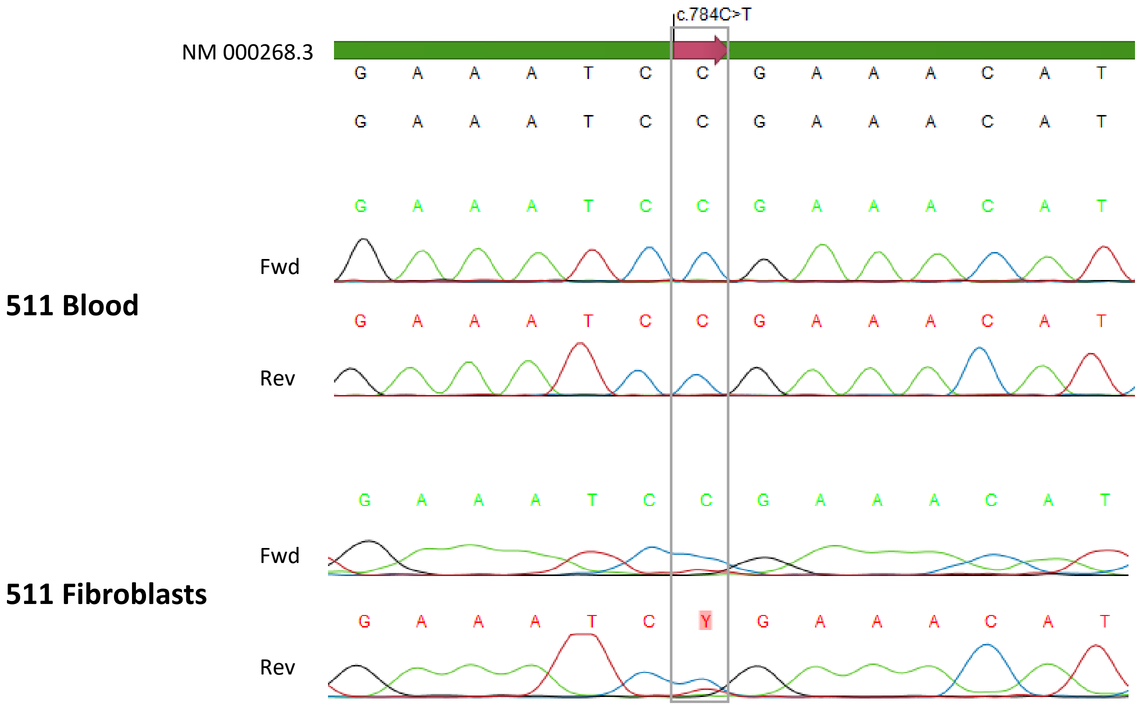

### Supplementary Table 1. NF2 Patients summary.

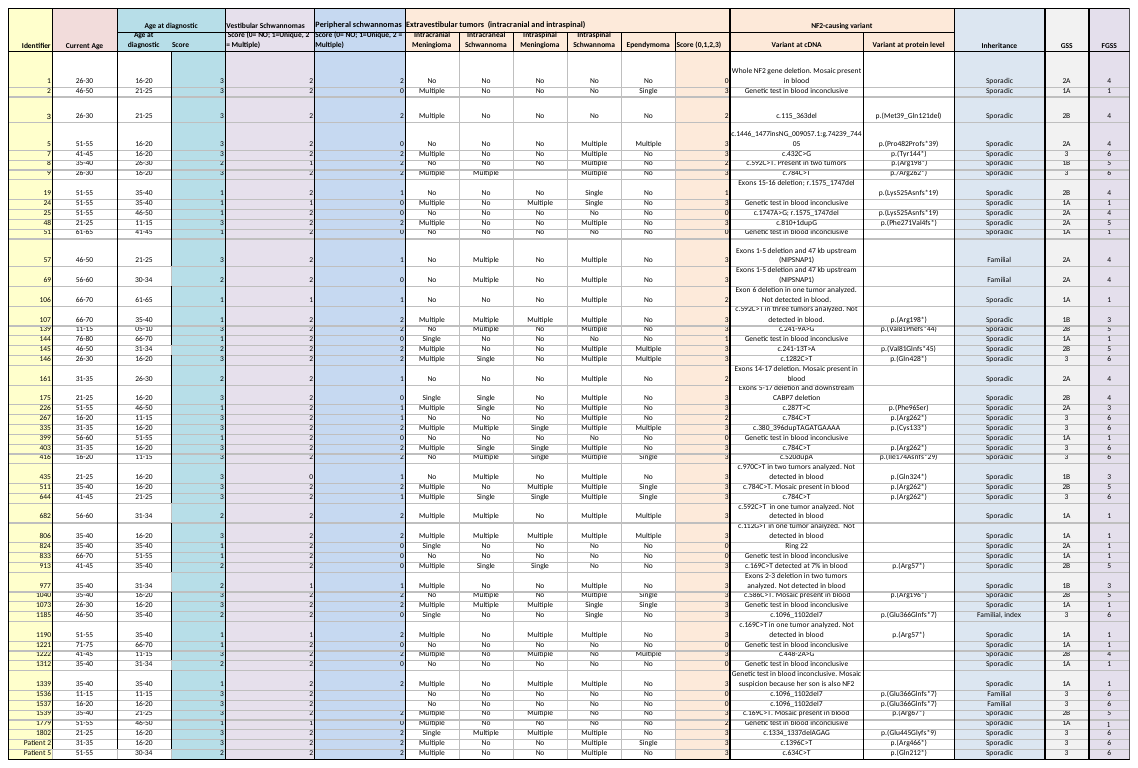
