## Supplementary Table 2. Major interventions in relation to Genetic Severity Score. for "Revisiting the UK Genetic Severity Score for NF2: a proposal for adding functional information"

| Genetic Severity | | | 1 Tissue Mosaic | 2A Mild | 2B Moderate | 3 Severe | Statistics |
|---|---|---|---|---|---|---|---|
| N (%) | Proportion of patients whithin | VS surgery | 7 (36,84%) | 9 (100%) | 5 (50%) | 10 (71,4%) | $X^2(1) = 4.08$, $p = 0.043$ |
| | | Non-VS intracranial surgery | 2 (10,5%) | 0 (0%) | 3 (30%) | 5 (35,7%) | $X^2(1) = 3.99$, $p = 0.045$ |
| | | Spinal surgery | 6 (31,58%) | 1 (11,11%) | 4 (40%) | 2 (14,3%) | $X^2(1) = 0.72$, $p = 0.39$ |
| | | Shunt surgery | 0 (0%) | 0 (0%) | 0 (0%) | 1 (7,1%) | $X^2(1) = 1.73$, $p = 0.18$ |
| | | Radiotherapy | 1 (5,26%) | 3 (33,33%) | 3 (30%) | 8 (57,1%) | $X^2(1) = 8.9$, $p = 0.002$ |
| | | Bevacizumab | 0 (0%) | 1 (11,11%) | 4 (40%) | 6 (42,9%) | $X^2(1) = 10.2$, $p = 0.001$ |
| | Total number of major interventions per person, grouped | 0 | 7 (36,84%) | 1 (11,11%) | 0 (0%) | 2 (14,3%) | $X^2(1) = 4.08$, $p = 0.043$ |
| | | 1 | 5 (26,31%) | 2 (22,22%) | 3 (30%) | 2 (14,3%) | $X^2(1) = 0.05$, $p = 0.82$ |
| | | 2 | 2 (10,52%) | 4 (44,44%) | 2 (20%) | 2 (14,3%) | $X^2(1) = 0$, $p = 1$ |
| | | 3 | 1 (5,26%) | 3 (33,33%) | 1 (10%) | 2 (14,3%) | $X^2(1) = 0.15$, $p = 0.69$ |
| | | 4 or more | 3 (15,79%) | 0 (0%) | 4 (40%) | 6 (42,9%) | $X^2(1) = 4.10$, $p = 0.042$ |
| Mean (SD) | Total number of major interventions per person, grouped | | 1,53 (1,91) | 1,90 (0,99) | 3,09 (2,34) | 3,79 (3,36) | $r_s(50) = 0.35$, $p = 0.01$ |
| | Number of total surgeries | | 1,53 (2,07) | 1,50 (0,71) | 1,91 (1,64) | 2,64 (2,98) | $r_s(50) = 0.18$, $p = 0.19$ |
| | Age at first radiotherapy session | | 23 | 37 (9,64) | 30,33 (6,66) | 24,29 (8,16) | $r_s(12) = -0.42$, $p = 0.13$ |
| | Age started bevacizumab | | - | 32 | 26,67 (14,57) | 23,67 (8,48) | $r_s(8) = -0.26$, $p = 0.46$ |
| | Age at first surgery | | 35,44 (13,36) | 29,78 (12,05) | 26,67 (7,57) | 20,80 (7,73) | $r_s(35) = -0.47$, $p = 0.003$ |
| | Age at first major intervention | | 34,20 (13,20 | 29,78 (12,05) | 25,55 (8,55) | 20,55 (7,89) | $r_s(39) = -0.47$, $p = 0.001$ |
| | Ratio of total number of major interventions to current age | | 0,03 (0,04) | 0,04 (0,02) | 0,08 (0,06) | 0,13 (0,11) | $r_s(50) = 0.49$, $p < 0.001$ |
