## Supplementary Table 3. NF2 Phenotype quantification. for "Revisiting the UK Genetic Severity Score for NF2: a proposal for adding functional information"

**Supplementary Table 3. Phenotype quantification**

| Phenotypic feature | | Value |
|---|---|---|
| Age at diagnosis | < 25 years | 3 |
| | < 35 years | 2 |
| | > 35 years | 1 |
| Vestibular Schwannoma | Unilateral | 1 |
| | Bilateral | 2 |
| Peripheral Schwannomas | Single | 1 |
| | Multiple | 2 |
| Extravestibular affection (Spinal/Cerebral) | Spinal and cerebral injury or spinal/cerebral different type of injury | 3 |
| | Cerebral or Spinal multiple injuries | 2 |
| | Single brain or spinal injury | 1 |
