## Supplementary Table 4. Revising pathogenicity associated to NF2 mutations according to GSS. Patients with an unexpected phenotype according to GSS are for "Revisiting the UK Genetic Severity Score for NF2: a proposal for adding functional information"

Suppplementary Table 4. Revising mutation associated pathogenicity according to GSS

| NF2-causing variant | | | GSS | Inheritance | Mosaicism | | Phenotype Quantification | Merlin NPE levels | pERK NPE levels | Reasoning |
|---|---|---|---|---|---|---|---|---|---|---|
| cDNA (NM_00268.3) | Exon | Predicted protein | | | % in blood | Only in tissue | | | | |
| c.169C>T & c.810+2T>A | exon 2; exon 8 | p.(Arg57*) & p.(?) | 1A | Sporadic | Not detected | One tumor analyzed | 3 | 3,66129319 | 4,0767212 | Mosaic patient. Very mild phenotype |
| c.1341-25_1377del62 + LOH | exon 13 | p.(?) | 1A | Sporadic | Not detected | One tumor analyzed | 3 | | | Mosaic patient. Very mild phenotype |
| Genetic test (NGS) in blood inconclusive | | | 1A | Sporadic | | | 3 | | | Mosaic patient. Very mild phenotype |
| Genetic test (NGS) in blood inconclusive | | | 1A | Sporadic | | | 3 | | | Mosaic patient. Very mild phenotype |
| Genetic test (NGS) in blood inconclusive | | | 1A | Sporadic | | | 4 | | | Mosaic patient. Very mild phenotype |
| Genetic test (NGS) in blood inconclusive | | | 1A | Sporadic | | | 4 | | | Mosaic patient. Very mild phenotype |
| Genetic test (NGS) in blood inconclusive | | | 1A | Sporadic | | | 4 | | | Mosaic patient. Very mild phenotype |
| Genetic test in blood inconclusive | | | 1A | Sporadic | | | 5 | | | Mosaic patient. Mild phenotype |
| exon 6 deletion + LOH | exon 6 | p.(Val173Glyfs*2) | 1A | Sporadic | Not detected | One tumor analyzed | 5 | | | Mosaic patient. Mild phenotype |
| c.169C>T + LOH | exon2 | p.(Arg57*) | 1A | Sporadic | Not detected | One tumor analyzed | 7 | | | **Mosaic patient. Unexpected mild-moderate phenotype** |
| Genetic test in blood inconclusive | | | 1A | Sporadic | | | 8 | | | **Mosaic patient. Unexpected moderate phenotype** |
| Genetic test (NGS) in blood inconclusive | | | 1A | Sporadic | | Mosaic suspicion be | 8 | | | **Mosaic patient. Unexpected moderate phenotype** |
| c.592C>T + LOH | exon 6 | p.(Arg198*) | 1A | Sporadic | 2% in fibroblas | One tumor analyzed | 9 | 3,57902164 | 9,08179524 | **Mosaic patient. Unexpected severe phenotype** |

| | | | | | | | | | | |
|---|---|---|---|---|---|---|---|---|---|---|
| c.112G>T + LOH | exon 1 | P.(Glu38*) | 1A | Sporadic | Not detected | One tumor analyzed | 10 | | | **Mosaic patient. Unexpected severe phenotype** |
| Genetic test (NGS) in blood inconclusive | | | 1A | Sporadic | | | 10 | | | **Mosaic patient. Unexpected severe phenotype** |
| exons 2-3 deletion | exon 2 & 3 | p.(?) | 1B | Sporadic | Not detected | Two tumors analyze | 7 | 2,28870175 | 3,0376054 | **Mosaic patient. Unexpected mild-moderate phenotype** |
| c.970C>T | exon 10 | p.(Gln324*) | 1B | Sporadic | Not detected | Two tumors analyze | 7 | 3,42502203 | 0,33804115 | **Mosaic patient. Unexpected mild-moderate phenotype** |
| c.592C>T | exon 6 | p.(Arg198*) | 1B | Sporadic | Not detected | Three tumors analyz | 8 | 4,58749871 | 4,55488321 | **Mosaic patient. Unexpected moderate phenotype** |
| c.592C>T | exon 6 | p.(Arg198*) | 1B | Sporadic | Not detected | Two tumors analyze | 7 | | | **Mosaic patient. Unexpected mild-moderate phenotype** |
| c.1747A>G; r.1575_1747del | exon 15 | p.(Lys525Asnfs*19) | 2A | Sporadic | | | 3 | 0,65585897 | 1,75666641 | Splicing variant at exon 15. Mild phenotype |
| Ring 22 | | | 2A | Sporadic | | | 3 | | | |
| exons 14-17 deletion | exons 14-17 | p.(?) | 2A | Sporadic | 20% (MLPA kit P044) | | 7 | 0,82372417 | 0,34586901 | **Large deletion excluding promoter in mosaicism. Unexpected mild-moderate phenotype. Merlin and pERK levels below threshold.** |
| c.287T>C | exon 3 | p.(Phe96Ser) | 2A | Sporadic | | | 7 | 2,18077529 | 2,23406472 | **Missense variant. Unexpected mild-moderate phenotype.** |
| Whole NF2 gene deletion in mosaicism | whole gene | | 2A | Sporadic | ~10% (MLPA kit P044) | | 7 | | | **Large deletion including promoter in mosaicism. Unexpected mild-moderate phenotype.** |
| exons 1-5 deletion and 47 kb upstream (NIPSNAP1) | exon 1-5 | Merlin synthesis altered | 2A | Familial | | | 7 | | | **Large deletion including promoter. Unexpected mild-moderate phenotype.** |
| c.1446_1477insNG_009057.1:g.74239_74405 | Deep intronic mutation between exons 13 - 14 | p.(Pro482Profs*39) | 2A | Sporadic | | | 8 | 0,41335449 | 4,7636402 | **Splicing variant at exon 14. Unexpected moderate phenotype. pERK levels above threshold.** |

| | | | | | | | | | | |
|---|---|---|---|---|---|---|---|---|---|---|
| exons 1-5 deletion and 47 kb upstream (NIPSNAP1) | exon 1-5 | Merlin synthesis altered | 2A | Familial | | | 9 | 1,68851611 | 5,7267768 | **Large deletion including promoter. Unexpected severe phenotype.** |
| c.810+1dupG | exon 8 | p.(Phe271Val4fs*) | 2A | Sporadic | | | 10 | 0,7651892 | 0,66179798 | **Splicing variant at exon 8. Unexpected severe phenotype. Merlin and pERK levels below threshold.** |
| exons 15-16 deletion; r.1575_1747del | exon15-16 | p.(Lys525Asnfs*19) | 2B | Sporadic | | | 5 | 0,98880623 | 5,64437297 | **Large deletion excluding promoter. Unexpected mild phenotype. At functional level this deletion alters exon 15 splicing. pERK levels are above threshold** |
| c.169C>T | exon 2 | p.(Arg57*) | 2B | Sporadic | Detected at 7% in blood (NGS) | | 7 | | | Nonsense variant at mosaicism. Mild-moderate phenotype |
| exons 5-17 deletion and downstream CABP7 deletion | exons 5-17 | p.(?) | 2B | Sporadic | | | 8 | 1,55439894 | 2,65118931 | Large deletion excluding promoter. Moderate phenotype. pERK levels are above threshold |
| c.115_363del | Exon 2&3 skipping | p.(Met39_Gln121del ) | 2B | Sporadic | | | 9 | 1,48734671 | 3,14955982 | Splicing variant at exon 2&3. Unexpected severe phenotype. pERK levels above threshold. |
| c.241-13T>A | exon 3 | p.(Val81Glnfs*45) | 2B | Sporadic | | | 9 | 2,06863943 | 1,09999436 | Splicing variant at exon 3. Unexpected severe phenotype. pERK levels below threshold. |
| c.241-9A>G | exon 3 | p.(Val81Phefs*44) | 2B | Sporadic | | | 10 | 1,57827175 | 2,46664781 | Splicing variant at exon 3. Unexpected severe phenotype. pERK levels below threshold. |
| c.784C>T | exon 8 | p.(Arg262*) | 2B | Sporadic | Detected at ~ 10% in fibroblasts, n[illegible] | | 10 | 2,56486969 | 0,81433044 | **Nonsense variant at mosaicism. Unexpected severe phenotype. pERK levels below threshold** |

| | | | | | | | | | | |
|---|---|---|---|---|---|---|---|---|---|---|
| c.169C>T | exon 2 | p.(Arg67*) | 2B | Sporadic | 5,6% in blood (NGS) | | 10 | 2,71920859 | 1,74534982 | **Nonsense variant at mosaicism. Unexpected severe phenotype. pERK levels below threshold** |
| c.586C>T | exon 6 | p.(Arg196*) | 2B | Sporadic | Detected at >20% in blood (externa | | 10 | | | **Nonsense variant at mosaicism. Unexpected severe phenotype** |
| c.448-2A>G | exon 5 | p.(?), splicing | 2B | Sporadic | | | 10 | | | Splicing variant at exon 3. Unexpected severe phenotype |
| c.1096_1102del7 | exon11 | p.(Glu366Glnfs*7) | 3 | Familial, index | | | 7 | 1,28967814 | 1,45881049 | Nonsense variant. Unexpected mild-moderate phenotype. Protein levels below threshold |
| c.784C>T | exon 8 | p.(Arg262*) | 3 | Sporadic | | | 8 | 1,64843072 | 1,12977722 | Nonsense variant. Unexpected moderate phenotype. Protein levels below threshold |
| c.784C>T | exon 8 | p.(Arg262*) | 3 | Sporadic | | | 9 | 1,72710678 | 0,74796226 | Nonsense variant. Severe phenotype. Protein levels below threshold |
| c.634C>T | exon 7 | p.(Gln212*) | 3 | Sporadic | | | 9 | | | Nonsense variant. Severe phenotype |
| c.380_396dupTAGATGAAAA | exon 4 | p.(Cys133*) | 3 | Sporadic | | | 10 | 0,6773879 | 0,81003198 | Nonsense variant. Severe phenotype. Protein levels below threshold |

| | | | | | | | | | | |
|---|---|---|---|---|---|---|---|---|---|---|
| c.1282C>T | exon 12 | p.(Gln428*) | 3 | Sporadic | | | 10 | 0,80455913 | 0,66749818 | Nonsense variant. Severe phenotype. Protein levels below threshold |
| c.432C>G | exon 4 | p.(Tyr144*) | 3 | Sporadic | | | 10 | 1,04771615 | 1,90213891 | Nonsense variant. Severe phenotype. Protein levels below threshold |
| c.784C>T | exon 8 | p./Arg262*) | 3 | Sporadic | | | 10 | 1,0694117 | 1,74014287 | Nonsense variant. Severe phenotype. Protein levels below threshold |
| c.784C>T | exon 8 | p.(Arg262*) | 3 | Sporadic | | | 10 | 1,17909458 | 0,32191149 | Nonsense variant. Severe phenotype. Protein levels below threshold |
| c.520dupA | exon 6 | p.(Ile174Asnfs*29) | 3 | Sporadic | | | 10 | 1,23139075 | 0,29106762 | Nonsense variant. Severe phenotype. Protein levels below threshold |
| c.1334_1337delAGAG | Small Deletion | p.(Glu445Glyfs*9) | 3 | Sporadic | | | 10 | | | Nonsense variant. Severe phenotype |
| c.1396C>T | exon 13 | p.(Arg466*) | 3 | Sporadic | | | 10 | | | Nonsense variant. Severe phenotype |
