## Supplementary Table 5. Demographic data according to Functional Genetic Severity Score. for "Revisiting the UK Genetic Severity Score for NF2: a proposal for adding functional information"

| Functional Genetic Severity Score | | | 1 | 2 | 3 | 4 | 5 | 6 | Correlation |
|---|---|---|---|---|---|---|---|---|---|
| N (% total) | Number of patients | | 16 (30,7%) | 0 | 4 (7,7%) | 10 (19,23%) | 8 (15,38%) | 14 (26,9%) | |
| N (% gender) | Gender | Male | 12 (36,4%) | | 3 (9,1%) | 6 (18,18%) | 3 (9%) | 9 (27,3%) | |
| | | Female | 4 (21,1%) | | 2 (10,5%) | 4 (21,1%) | 4 (21,1%) | 5 (26,3%) | $X^2(4) = 1.81, p = 0.77$ |
| Mean (SD) | Age at diagnosis | | 40,94 (16,11) | | 36,6 (12,99) | 23 (7,37) | 24 (12,90) | 18,79 (6,5) | $r_s(50) = -0.64, p < 0.001$ |
| | Current age | | 52,25 (15,56) | | 46,8 (18,43) | 41,56 (12,76) | 36,88 (12,91) | 30,79 (12,07) | $r_s(50) = -0.54, p < 0.001$ |
| | Years since diagnosis | | 11,25 (6,86) | | 10,2 (12,21) | 18,56 (12,09) | 12,88 (6,69) | 12 (8,26) | $r_s(50) = 0.04, p = 0.75$ |
| | Age at NF2-related death | | | | | 53 | 40 | 42 | |
| N (% score category) | NF2-related deaths | | | | | 1 (10%) | 1 (12,5%) | 1 (7,2%) | |
| | Familial NF2 | | 0 (0%) | | 1 (20%) | 3 (30%) | 0 (0%) | 3 (21,4%) | |
| | Sporadic NF2 | | 16 (100%) | | 4 (80%) | 7 (70%) | 7 (87,5%) | 11 (78,6%) | X2(4) = 7.73, p = 0.10 |
