## Supplementary Table 6. Tumor burden, presence of ocular features and hearing outcome according to Functional Genetic Severity Score for "Revisiting the UK Genetic Severity Score for NF2: a proposal for adding functional information"

| **Functional Genetic Severity Score** | | | | **1** | **2** | **3** | **4** | **5** | **6** | **Statistics** |
|---|---|---|---|---|---|---|---|---|---|---|
| N (% total) | Number of patients | | | 16 | 0 | 4 | 10 | 8 | 14 | |
| Tumor load | N (%) | Bilateral VS | | 11 (68,7%) | 0 (0%) | 3 (75%) | 10 (100%) | 8 (100%) | 14 (100%) | $X^2(1) = 8.9, p = 0.002$ |
| | | Unilateral VS | | 5 (31,2%) | 0 (0%) | 1 (25%) | 0 (0%) | 0 (0%) | 0 (0%) | $X^2(1) = 8.9, p = 0.002$ |
| | | Intracranial meningioma | | 9 (56,2%) | 0 (0%) | 3 (75%) | 3 (30%) | 5 (62,5%) | 10 (71,4%) | $X^2(1) = 0.35, p = 0.55$ |
| | | Intracranial Schwannoma | | 3 (18,7%) | 0 (0%) | 3 (75%) | 3 (30%) | 3 (37,5%) | 7 (50%) | $X^2(1) = 1.8, p = 0.17$ |
| | | Spinal meningioma | | 5 (31,2%) | 0 (0%) | 1 (25%) | 1 (10%) | 3 (37,5%) | 5 (45,5%) | $X^2(1) = 0.10, p = 0.74$ |
| | | Spinal Schwannoma | | 8 (50%) | 0 (0%) | 4 (100%) | 6 (60%) | 6 (75%) | 11 (100%) | $X^2(1) = 1.7, p = 0.17$ |
| | | Spinal ependymoma | | 4 (25%) | 0 (0%) | 0 (0%) | 2 (20%) | 3 (37,5%) | 5 (45,5%) | $X^2(1) = 0.8, p = 0.35$ |
| Ocular Features | N (%) | Epiretinal membranes | | 0 (0%) | 0 (0%) | 0 (0%) | 0 (0%) | 0 (0%) | 0 (0%) | NA |
| | | Cataract | | 3 (37,5%) | 0 (0%) | 1 (50%) | 3 (30%) | 1 (12,5%) | 3 (42,9%) | $X^2(1) = 0, p = 1$ |
| | | Combined hamartoma | | 0 (0%) | 0 (0%) | 0 (0%) | 0 (0%) | 0 (0%) | 2 (28,6%) | $X^2(1) = 3.1, p = 0.07$ |
| | | Optic nerve meningioma | | 0 (0%) | 0 (0%) | 0 (0%) | 0 (0%) | 1 (12,5%) | 0 (0%) | $X^2(1) = 0.3, p = 0.53$ |
| | Mean (SD) | Total eye features | | | 0 (0%) | | | | | $r_s(49) = 0.06, p = 0.67$ |
| Hearing Outcomes | N (%) | Hearing grade | 1 | 12 (75%) | 0 (0%) | 4 (100%) | 3 (30%) | 3 (37,5%) | 6 (42,9%) | $X^2(1) = 5.4, p = 0.019$ |
| | | | 2 | 1 (6,2%) | 0 (0%) | 0 (0%) | 0 (0%) | 0 (0%) | 0 (0%) | $X^2(1) = 1.5, p = 0.20$ |
| | | 3 o 4 | | 0 (0%) | 0 (0%) | 0 (0%) | 0 (0%) | 2 (25%) | 1 (7,1%) | $X^2(1) = 2.1, p = 0.14$ |
| | | | 5 | 2 (12,5%) | 0 (0%) | 0 (0%) | 3 (30%) | 1 (12,5%) | 4 (28,6%) | $X^2(1) = 1.6, p = 0.20$ |
| | | | 6 | 2 (12,5%) | 0 (0%) | 0 (0%) | 4 (40%) | 2 (25%) | 3 (21,4%) | $X^2(1) = 0.6, p = 0.42$ |
| | Mean (SD) | Age of loss of useful hearing | | 48,31 (17,16) | 0 (0%) | 46,8 (18,43) | 32,38 (8,43) | 29,38 (11,89) | 26,23 (12,58) | $r_s(48) = -0.58, p < 0.001$ |
| Cutaneus manifestations | N (%) | | | 4 (25%) | 0 (0%) | 0 (0%) | 0 (0%) | 4 (50%) | 9 (75%) | $X^2(1) = 6.2, p = 0.012$ |

* For group 1 and 6, only 4 out 16 and 11 out of 14 patients, respectively, have been tested by a spinal magnetic resonance
