## Supplementary Table 7. Major interventions in relation to Functional Genetic Severity Score. for "Revisiting the UK Genetic Severity Score for NF2: a proposal for adding functional information"

| **Functional Genetic Severity Score** | | | **1** | **2** | **3** | **4** | **5** | **6** | **Statistics** |
|---|---|---|---|---|---|---|---|---|---|
| N (%) | Proportion of patients | VS surgery | 6 (37,5%) | | 3 (60%) | 9 (90%) | 3 (37,5%) | 10 (71,4%) | $X^2(1) = 2.16$, p = 0.14 |
| | | Non-VS intracranial surgery | 1 (6,3%) | | 1 (20%) | 0 (0%) | 3 (37,5%) | 5 (35,7%) | $X^2(1) = 4.5$, p = 0.032 |
| | | Spinal surgery | 3 (18,8%) | | 4 (80%) | 1 (10%) | 3 (37,5%) | 2 (14,3%) | $X^2(1) = 0.4$, p = 0.51 |
| | | Shunt surgery | 0 (0%) | | 0 (0%) | 0 (0%) | 0 (0%) | 1 (7,1%) | $X^2(1) = 1.5$, p = 0.21 |
| | | Radiotherapy | 1 (6,3%) | | 0 (0%) | 3 (30%) | 3 (37,5%) | 8 (57,1%) | $X^2(1) = 10.1$, p = 0.001 |
| | | Bevacizumab | 0 (0%) | | 0 (0%) | 2 (20%) | 3 (37,5%) | 6 (42,9%) | $X^2(1) = 9.6$, p = 0.002 |
| | Total number of major interventions per person, grouped | 0 | 8 (50%) | | 0 (0%) | 0 (0%) | 0 (0%) | 2 (14,3%) | $X^2(1) = 7.1$, p = 0.007 |
| | | 1 | 3 (18,8%) | | 3 (60%) | 2 (20%) | 2 (25%) | 2 (14,3%) | $X^2(1) = 0.4$, p = 0.51 |
| | | 2 | 2 (12,5%) | | 1 (20%) | 4 (40%) | 1 (12,5%) | 2 (14,3%) | $X^2(1) = 0.03$, p = 0.86 |
| | | 3 | 1 (6,3%) | | 0 (0%) | 4 (40%) | 0 (0%) | 2 (14,3%) | $X^2(1) = 0.22$, p = 0.63 |
| | | 4 or more | 2 (12,5%) | | 1 (20%) | 0 (0%) | 4 (50%) | 6 (42,9%) | $X^2(1) = 4.5$, p = 0.033 |
| Mean (SD) | Total number of major interventions per person, grouped | | 1,25 (1,77) | | 2 (1,73) | 2,22 (0,83) | 3,63 (2,50) | 3,79 (3,36) | $r_s(50) = 0.39$, p = 0.003 |
| | Number of total surgeries | | 1,19 (1,80) | | 2,2 (2,17) | 1,67 (0,50) | 2,13 (1,89) | 2,64 (2,98) | $r_s(50) = 0.22$, p = 0.11 |
| | Age at first radiotherapy session | | 23 | | 0 | 37 (9,64) | 30,33 (6,66) | 24,29 (8,16) | $r_s(12) = -0.42$, p = 0.13 |
| | Age started bevacizumab | | 0 | | 0 | 32 | 26,67 (14,57) | 23,67 (8,48) | $r_s(8) = -0.26$, p = 0.46 |
| | Age at first surgery | | 36,71 (13,90) | | 34,8 (10,62) | 25,67 (7,55) | 27,50 (11,93) | 20,80 (7,73) | $r_s(35) = -0.51$, p = 0.001 |
| | Age at first major intervention | | 35,00 (13,75) | | 34,8 (10,62) | 25,67 (7,55) | 25,75 (11,93) | 20,55 (7,89) | $r_s(39) = -0.51$, p < 0.001 |
| | Ratio of total number of major interventions to current age | | 0,03 (0,04) | | 0,04 (0,02) | 0,05 (0,02) | 0,10 (0,06) | 0,13 (0,11) | $r_s(50) = 0.54$, p < 0.001 |
