## Supplementary Table 8. Intragroup Variability of Merlin, pERK, age at diagnosis and age at hearing loss. for "Revisiting the UK Genetic Severity Score for NF2: a proposal for adding functional information"

| Score | GSS | | | | | FGSS | | | | | |
|---|---|---|---|---|---|---|---|---|---|---|---|
| **Group/Class** | **1** | **2A** | **2B** | **3** | **Mean** | **1** | **3** | **4** | **5** | **6** | **Mean** |
| **Merlin Protien Levels (σ)** | 3,11 | 0,54 | 0,29 | 0,12 | 1,02 | 1,76 | 2,01 | 0,27 | 0,55 | 0,12 | 0,94 |
| **pERK Protien Levels(σ)** | 6,88 | 3,99 | 5,43 | 0,54 | 4,21 | 3,54 | 3,27 | 2,01 | 0,44 | 0,34 | 1,92 |
| **Age at diagnosis (σ)** | 216,97 | 169,78 | 99,46 | 33,27 | 129,87 | 202,77 | 149,08 | 54,29 | 182,25 | 35,62 | 124,80 |
| **Age at hearing loss (σ)** | 324,72 | 144 | 110,88 | 148,1 | 181,925 | 262,11 | 365,19 | 71,47 | 148,35 | 117,93 | 193,01 |
