## Supplementary Table 9. A) FGSS and NF2 disease-causing variant regression model. B) FGSS and NF2 disease-causing variant regression model. for "Revisiting the UK Genetic Severity Score for NF2: a proposal for adding functional information"

**Suppplementary Table 9A. FGSS and NF2 disease-causing variant regression model**

| | Model 1 (N=52) | | | | Model 2 (N=27) | | | |
|---|---|---|---|---|---|---|---|---|
| | **beta** | **se** | **t** | **p** | **beta** | **se** | **t** | **p** |
| **Intercept** | 5062 | 0.519 | 9739 | 7.46e-13 | 5141 | 18612 | 2762 | 0.012 |
| **Type 3** | 2187 | 1162 | 1882 | 0.066 | | | | |
| **Type 4** | 3080 | 0.942 | 3269 | 0.002 | 2419 | 14691 | 1647 | 0.116 |
| **Type 5** | 2837 | 0.838 | 3385 | 0.001 | 2035 | 13829 | 1472 | 0.157 |
| **Type 6** | 3804 | 0.747 | 5091 | 6.18e-06 | 3352 | 13817 | 2426 | 0.025 |
| **pERK** | | | | | -0.135 | 0.241 | -0.562 | 0.580 |
| **NF2** | | | | | 0.786 | 0.499 | 1574 | 0.132 |
| | | | $R^2$=0.381 | 0.0001 | | | $R^2$=0.143 | 0.161 |

The outcome in both models is FGSS. The patients in Model 2 only have mutations of type 3, 4, 5 and 6. Then, the base category of this model is Type 3.

**Suppplementary Table 9B. GSS and NF2 disease-causing variant regression model**

| | Model GSS (N=52) | | | |
|---|---|---|---|---|
| | **beta** | **se** | **t** | **p** |
| **Intercept** | 5428 | 0.565 | 9603 | 2.3e-12 |
| **1B** | 1904 | 1345 | 1415 | 0.163 |
| **2A** | 1349 | 0.903 | 1493 | 0.142 |
| **2B** | 3207 | 0.852 | 3764 | 0.0004 |
| **3** | 3238 | 0.832 | 3891 | 0.0003 |
| | | | $R^2$=0.322 | 0.001 |

The outcome in the model is GSS.
