## Supplementary Materials and Methods for "Revisiting the UK Genetic Severity Score for NF2: a proposal for adding functional information"

**Supplementary Methods and Materials:**

**Genetic NF2 test:** Genetic test was performed using the customized I2HCP panel (Castellanos et al 2017) in blood or tissue when available and validated by Sanger sequencing in all primary cultures established. In mosaic patients, a deep sequencing with a >700 depth of coverage was performed in primary culture fibroblasts. Only variants detected above 5% of the cells were considered.

**Functional assay, Western Blot analysis:** Merlin’s downstream pathways were analyzed in 27 out of 30 NF2 patients included in the validation of the Genetic Severity Score. We obtained cultured fibroblasts from skin biopsies that were processes as described in [29]. 50 µg of protein extracted from these primary cultures at 50%, 80% and 100% of confluence was load to SDS-PAGE (150V) and transferred onto PVDF membranes (1 hour 350 mA at 4^o^C). Odyssey Blocking Buffer TBS (LI-COR) was used to block the resulting membranes. Three primary cultures from healthy donors were included as control samples (Ctrl1, Ctrl2 and Ctrl3). When grouping for the statistical studies, it has been taken into account the control healthy donors and the mosaics as a control group, since none of these show the constitutional *NF2* mutation in the analyzed tissue. Primary antibodies were incubated at 4^o^C overnight. Membranes were later incubated with IRDye 680LT and IRDye 800CW secondary antibodies (1:1000 dilution, LI-COR) for 1h at room temperature and scanned and analyzed using the Odyssey Infrared Imaging System (LI-COR). The statistical threshold that allows the differentiation between healthy controls or mosaic patients and the patients in group 3 (severe) is established through the ±2SD limits method.

**Primary antibodies:** **α-pERK**: phospho-p44/42; APK (Erk1/2) (Thr202/Tyr204) (E10) #9106 α-mouse (Cell Signaling Technology), **α-ERK**: p44/42; APK (Erk1/2) Antibody #9102 – α – rabbit (Cell Signaling Technology, **α-pAKT:** Phospho Akt (Ser 473) (587F11) Mouse ab #4051 α-mouse (Cell Signaling Technology), **α-AKT** Akt Antibody #9272– α –rabbit (Cell Signaling Technology, **α-pNF2:** Phospho NF2/Merlin (phospho S518) antibody (ab131473) α-rabbit (Abcam), **α-NF2:** NF2/Merlin antibody (ab88957) α-mouse (Abcam), **α -PAK1:** Anti-PAK1 antibody (ab154284) α-rabbit (Abcam), **α -RAC1:** Anti-RAC1 antibody (ab33186) α-mouse (Abcam), **α-pYAP1**: anti-YAP1 (phospho S127) antibody [EP1675Y] (ab76252) – α – rabbit (Abcam), **α-YAP1**: anti-YAP1 antibody (ab56701) – α – mouse (Abcam), **α-pPKA:** Phospho PKA. Anti PKA alpha+beta (phospho T197) antibody (ab5815) – α – rabbit (Abcam), **α-PKA:** PKA C alpha beta antibody Clone #515741 (MAB5908) – α – mouse (Abcam). **α-vinculin:** anti-vinculin antibody [EPR8185] (ab129002) – α – rabbit (Abcam) was used to normalize protein expression among samples

**NF2 phenotype quantification**: Clinical phenotype was recorded as follows: age of diagnosis was rated as 3 if it was before 25 years, 2 between 25 and 35 years and 1 if patient was diagnosed after 35; the presence of peripheral and vestibular schwannomas were punctuate as 0, 1 or 2 if they were absent, unique or multiple, while extravestibular lesions were score between 1 if we detected single intracranial or intra-spinal lesion, 2 if the patients showed multiple intracranial or intra-spinal schwannomas or meningiomas, or 3 when patient developed multiple intra-spinal or intracerebral different lesion types or multiple intra-spinal and intracerebral schwannomas or meningiomas (Supplementary Table 3).

**NF2 pathogenic variant score**: Truncating mutations in exons 2-13 were scored 6, missense and in frame deletions as 3, presumed mosaics or patients carrying a Ring22 as 1. Based in [7,10,14,41], the predicted effect of *NF2* mutation in Merlin’s function, and our observations in the phenotype shown by patients harboring mutations in the most terminal region of the gene and the results from the pERK functional analysis, we decided to rate differently when mutations are located in first exons, as Merlin could start its translation using another methionine, or if the variant does not alter *NF2* reading frame because some Merlin residual function could be maintained. In addition, it was also considered when the variant affects exon 14 or the followings due to its association to good prognosis (Supplementary Table 4) [7,8]. In addition, without more evidences, we scored all missense variants as 3, since in our functional assays and as published before [42], missense mutations result in the quantitative loss of merlin proteins, and we assume they might minimally affect the intrinsic protein function unless when altering specific positions, which would be rated differently once demonstrated the association to a severe phenotype. In contrast to GSS, we scored as less severe those splicing and copy number alterations (CNA) at exon level that do not alter the reading frame in comparison as those generating a frameshift alteration assuming that in frame alterations could generate an hypomorphic Merlin, but we do not consider differently when variants are located at exons 1-7 or 8-13. We also accounted generalized mosaicism if *NF2* variant was detected in blood or unaffected tissue differently from tissue mosaicism if it was detected only in two independent tumors, since the affection degree could be different. Finally, patients with an inconclusive blood test without any tumor analyzed were considered presumed tissue mosaics and scored as 1 (Table 3).

**Statistics:** All statistical analyses of the validation of the GSS were reproduced from the study of Halliday et al. Chi-square tests were performed to associate NF2 mutations with categorical phenotypical variables (i.e tumor load, ocular features, hearing outcomes and major interventions of the patients). Kruskal-Wallis test were performed to associate NF2 mutations with qualitative phenotypical variables transformed to categorical variables (i.e. age at diagnosis). Principal component analysis (PCA) of age at diagnosis, extravestibular affection, vestibular and peripheral schwannomas was performed to explore variability of NF2 mutations of patients. In order to study differences between protein expression levels in the functional assay, Kruskal-Wallis and Mann-Whitney U statistical tests were performed among genetic severity groups and mutation classes. Fibroblast protein levels from healthy controls have been included in group 1 for the statistics analysis. Analysis of variance (ANOVA) of FGSS and NF2 mutations was performed to test differences of severity between NF2 mutations groups. Shapiro Wilk and Levene tests were applied for testing Normality of residuals and homoscedasticity of groups respectively. Backward stepwise regression analysis was performed to evaluate the contribution of Merlin and pERK levels in the ANOVA model. Statistical analyses were performed using R software.
